# AnthropoAge, a novel approach to integrate body composition into the estimation of biological age

**DOI:** 10.1101/2021.09.23.21263703

**Authors:** Carlos A. Fermín-Martínez, Alejandro Márquez-Salinas, Enrique C. Guerra, Lilian Zavala-Romero, Neftali Eduardo Antonio-Villa, Luisa Fernández-Chirino, Eduardo Sandoval-Colin, Daphne Abigail Barquera-Guevara, Alejandro Campos Muñoz, Arsenio Vargas-Vázquez, César Daniel Paz-Cabrera, Daniel Ramírez-García, Luis Miguel Gutiérrez-Robledo, Omar Yaxmehen Bello-Chavolla

## Abstract

Aging is believed to occur across multiple domains, one of which is body composition; however, attempts to integrate it into biological age (BA) have been limited. Here, we consider the sex-dependent role of anthropometry for prediction of 10-year all-cause mortality using data from 18,794 NHANES participants to generate and validate a new BA metric. Our data-driven approach pointed to sex-specific contributors for BA estimation: WHtR, arm and thigh circumferences for men; weight, WHtR, thigh circumference, subscapular and triceps skinfolds for women. We used these measurements to generate AnthropoAge, which predicted all-cause mortality (AUROC 0.876, 95%CI 0.864-0.887) and cause-specific mortality independently of race, sex, and comorbidities; AnthropoAge was a better predictor than PhenoAge for cerebrovascular, Alzheimer and COPD mortality. A metric of age acceleration was also derived and used to assess sexual dimorphisms linked to accelerated aging, where women had an increase in overall body mass plus an important subcutaneous to visceral fat redistribution, and men displayed a marked decrease in fat and muscle mass. Finally, we showed that consideration of multiple BA metrics may identify unique aging trajectories with increased mortality (HR for multidomain acceleration 2.43, 95%CI 2.25-2.62) and comorbidity profiles. A simplified version of AnthropoAge (S-AnthropoAge) was generated using only BMI and WHtR, all results were preserved using this metric. In conclusion, AnthropoAge is a useful proxy of BA that captures cause-specific mortality and sex dimorphisms in body composition, and it could be used for future multidomain assessments of aging to better characterize the heterogeneity of this phenomenon.

**GRAPHICAL ABSTRACT:** 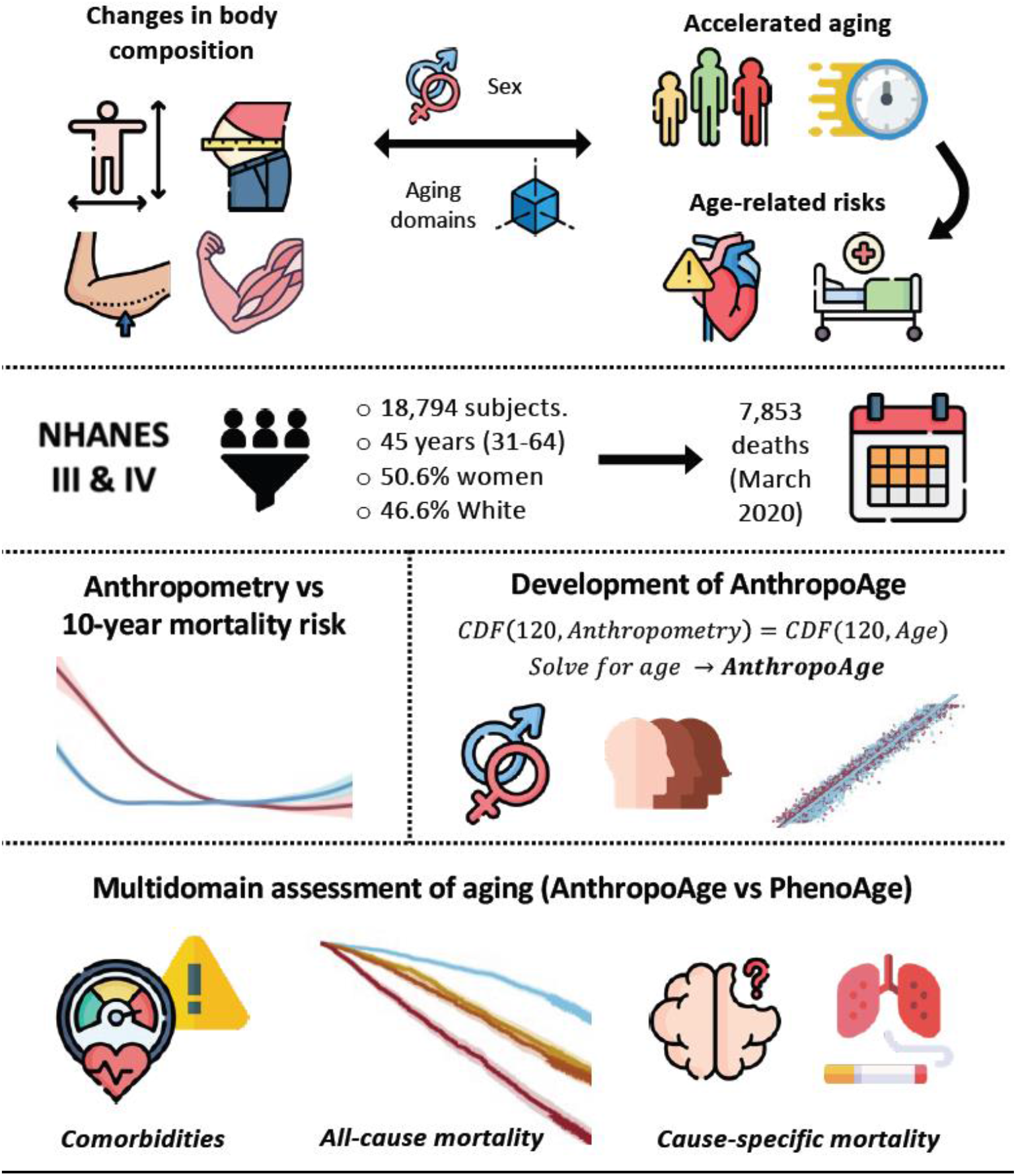

## INTRODUCTION

Aging is a complex phenomenon that researchers have been attempting to characterize for decades, and it is well known that chronological age (CA) does not fully capture its heterogeneity (Kennedy et al., 2014). Methods developed to quantify aging rates need to address the most common physiological changes that occur with aging and capture the variability of individual aging trajectories (López-Otín et al., 2013; Sebastiani et al., 2017). The concept of biological age (BA) goes beyond CA and refers to the underlying processes that modify the susceptibility for development of age-related diseases, disability, and functional impairment, ultimately increasing mortality risk (Belsky et al., 2015; Levine, 2013). Besides calculations derived from epigenetic and omics-based markers, previous efforts have used easily-accessible clinical tools to develop BA estimations, such as Phenotypic Age (PhenoAge), which uses CA and nine blood biomarkers (albumin, creatinine, glucose, C-reactive protein [CRP], mean corpuscular volume [MCV], red blood cell distribution width [RDW], alkaline phosphatase [ALP], white blood cell count [WBC] and lymphocyte percentage) for prediction of 10-year mortality risk (Levine et al., 2018; Liu et al., 2018). Aging is known to be a systemic process, but it is also believed to take place across domains that act in synergy to produce functional changes (Margolick & Ferrucci, 2015; P. -L. Kuo et al., 2020). In line with this idea, it has been shown that some BA estimations tend to be poorly correlated with each other (Li et al., 2020; Earls et al., 2019), while other studies have described BA metrics that capture distinct aspects of aging with unique genetic determinants (C. Kuo et al., 2021). We expect that a broader assessment of aging may provide a more comprehensive overview of its mechanisms, including additional aging domains not previously accounted for in BA estimations (Rivero-Segura et al., 2020). Changes in anthropometry and body composition have been one of the most extensively documented aspects of aging. Overall, aging is characterized by a linear decline in body length, while body mass initially increases in early and middle adulthood but starts decreasing in older individuals (Fernihough & McGovern, 2015; P. -L. Kuo et al., 2020), a reduction in muscle mass that accelerates at older ages also occurs, as well as a gradual increase in fat mass that is accompanied by a redistribution of adipose tissue from subcutaneous to visceral depots (Wilkinson et al., 2018; Tchernof & Després, 2013); in general, this shift is reflected by a shortening and broadening of the upper body in relation to the lower body (Frenzel et al., 2020). Interestingly, these changes take place at different rates and magnitudes for men and women; to be specific, men appear to have a steeper decline in muscle mass, and the redistribution of adipose tissue is more prominent in women (P. -L. Kuo et al., 2020; Frenzel et al., 2020). These modifications may reflect changes in lifestyle, but they can also be attributed to physiological alterations that occur with aging. Particularly, decline in lean mass can be explained by impaired responses to anabolic stimuli due to systemic inflammation, intramuscular lipid accumulation, stem cell exhaustion, or mitochondrial dysfunction, resulting in a poor regenerative potential of skeletal muscle (López-Otín et al., 2013; Wilkinson et al., 2018). On the other hand, neuroendocrine disturbances such as decrease in sexual hormones (specially in post-menopausal women), insulin resistance and systemic inflammation can hinder the capacity of the subcutaneous adipose tissue to store lipids, resulting in a disproportionate accumulation of visceral and ectopic fat (Tchernof & Després, 2013). Thus, changes in body composition can interact with each other, as well as with other genetic and environmental factors to produce unique age-related risk phenotypes that likely capture alterations in body homeostasis, including frailty, sarcopenic obesity, osteosarcopenia and metabolically unhealthy obesity (April-Sanders & Rodriguez, 2021; Atkins & Wannamathee, 2020; Fried et al., 2021). With all this, it is unsurprising that body composition is often recognized as one of the main domains in which aging takes place (P. -L. Kuo et al., 2020). However, despite the overwhelming evidence supporting the contribution of body composition and anthropometry to predict age and age-related outcomes, their proper implementation in the development of BA estimations has been limited (Kang et al., 2012; Negasheva et al., 2017). Here, we hypothesize that the integration of sex-based differences in multiple anthropometric measurements would result in the development of a reliable and easily implementable BA estimation that captures the body composition domain of aging. Moreover, we use a combination of PhenoAge and our new metric to investigate whether the simultaneous assessment of more than one domain of aging would identify unique trajectories with differences in mortality risk and body composition patterns. To test these hypotheses, we use anthropometric, laboratory and mortality data from two independent cycles of the National Health Examination Survey, with NHANES-III (1988-1994) used as the training cohort and NHANES-IV (1999-2008) as the validation cohort.

## RESULTS

### Anthropometric measurements included

For the purposes of our study, we divided anthropometric measurements available in all NHANES cycles into five groups that represent the property of body composition that they likely capture. 1) Body length: height, upper arm length and upper leg length. 2) Body mass: weight and body mass index (BMI). 3) Visceral adiposity: waist circumference and waist-to-height ratio (WHtR). 4) Subcutaneous adiposity: subscapular and triceps skinfolds. 5) Primarily lean mass: mid-upper arm circumference and mid-thigh circumference. (See **Supplementary Table 1**).

**TABLE 1.**
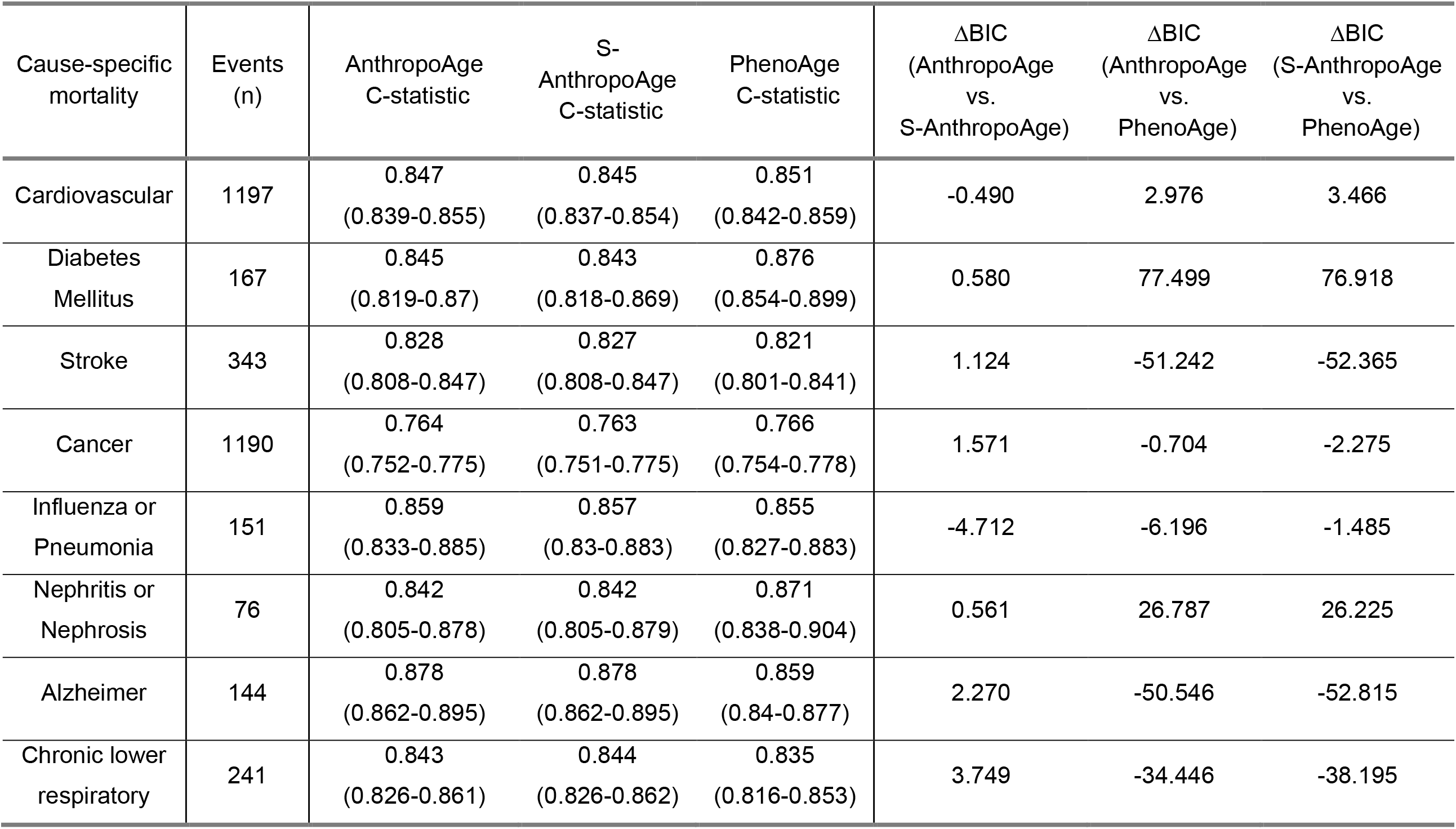
Performance of Fine & Gray semiparametric competitive risk regression models to evaluate cause-specific mortality using AnthropoAge, S-AnthropoAge and PhenoAge. We show the C-statistic (95% confidence interval) and the difference in the Bayesian Information Criterion (ΔBIC = BIC_Model-1_ - BIC_Model-2_) to contrast model predictions. ΔBIC < -2 indicates that model 1 is better. ΔBIC > 2 indicates that model 2 is better. All models are adjusted for age, sex, race/ethnicity, and number of comorbidities. Hazard ratios with 95% confidence intervals and standard errors for all models are available within supplementary material.

| Cause-specific mortality | Events (n) | AnthropoAge C-statistic | S-AnthropoAge C-statistic | PhenoAge C-statistic | ΔBIC (AnthropoAge vs. S-AnthropoAge) | ΔBIC (AnthropoAge vs. PhenoAge) | ΔBIC (S-AnthropoAge vs. PhenoAge) |
| --- | --- | --- | --- | --- | --- | --- | --- |
| Cardiovascular | 1197 | 0.847<br>(0.839-0.855) | 0.845<br>(0.837-0.854) | 0.851<br>(0.842-0.859) | -0.490 | 2.976 | 3.466 |
| Diabetes Mellitus | 167 | 0.845<br>(0.819-0.87) | 0.843<br>(0.818-0.869) | 0.876<br>(0.854-0.899) | 0.580 | 77.499 | 76.918 |
| Stroke | 343 | 0.828<br>(0.808-0.847) | 0.827<br>(0.808-0.847) | 0.821<br>(0.801-0.841) | 1.124 | -51.242 | -52.365 |
| Cancer | 1190 | 0.764<br>(0.752-0.775) | 0.763<br>(0.751-0.775) | 0.766<br>(0.754-0.778) | 1.571 | -0.704 | -2.275 |
| Influenza or Pneumonia | 151 | 0.859<br>(0.833-0.885) | 0.857<br>(0.83-0.883) | 0.855<br>(0.827-0.883) | -4.712 | -6.196 | -1.485 |
| Nephritis or Nephrosis | 76 | 0.842<br>(0.805-0.878) | 0.842<br>(0.805-0.879) | 0.871<br>(0.838-0.904) | 0.561 | 26.787 | 26.225 |
| Alzheimer | 144 | 0.878<br>(0.862-0.895) | 0.878<br>(0.862-0.895) | 0.859<br>(0.84-0.877) | 2.270 | -50.546 | -52.815 |
| Chronic lower respiratory | 241 | 0.843<br>(0.826-0.861) | 0.844<br>(0.826-0.862) | 0.835<br>(0.816-0.853) | 3.749 | -34.446 | -38.195 |

### Study population

We included only participants ≥20 years old with complete anthropometric and mortality data, resulting in a total of 18,794 subjects (NHANES-III: 11,774, NHANES-IV: 7,020). A flowchart diagram of participant selection for each analysis is shown in **Figure 1**, and a comprehensive overview of our study population is available in **Supplementary Table 2**. Briefly, among participants from both NHANES-III and NHANES-IV, 9,505 were female (50.6%) with a median CA of 45 years (IQR 31-64 years) and a median PhenoAge of 56.8 years (IQR 42.6-76.2 years), most participants self-identified as non-Hispanic White (8,761, 46.6%), followed by Mexican American (4,760, 25.3%), and non-Hispanic Black (4,246, 22.6%). Regarding medical history, 9,268 participants (49.3%) had at least one comorbidity, with hypertension (26.6%) being the most frequent. When assessing the distribution of anthropometric measurements, we found that men had greater values than women for all measurements except for WHtR, subscapular skinfold and triceps skinfold (**Figure 2**). We obtained all-cause mortality follow-up data using information from the National Death Index through March 2020, and a total of 7,853 deaths (31.1%) were recorded, with a mean follow-up of 191 months (IQR 131-288 months). Finally, we compared data from NHANES-III vs NHANES-IV and observed no significant differences in CA; however, median PhenoAge values were higher in NHANES-III (57.9 years [IQR 43.8-78.1] vs. 55.1 years [IQR 40.6-72.6], p<0.001). We observed that all anthropometric variables significantly differed between individuals from both cohorts except for subscapular skinfold for men and BMI for women.

**FIGURE 1.**
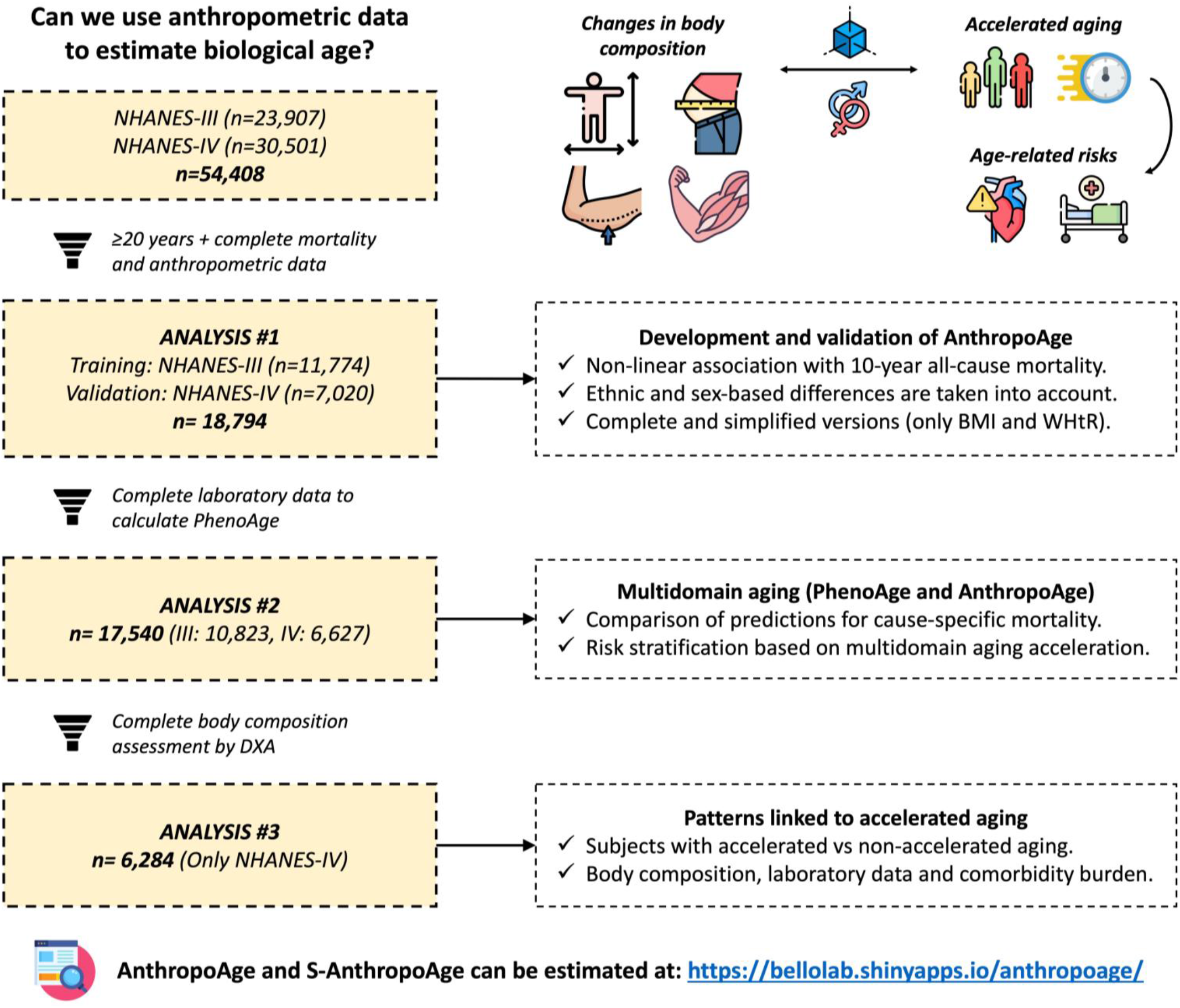
Flowchart diagram of participant selection from the National Health and Nutrition Examination Survey (NHANES) along with the key points from each analysis in our study. A brief conceptual framework is also provided at the top, illustrating how changes in body composition can modify aging rates, ultimately leading to accelerated aging and poor age-related outcomes, and this relationship is strongly influenced by sex and by changes in other domains of aging.

**FIGURE 2.**
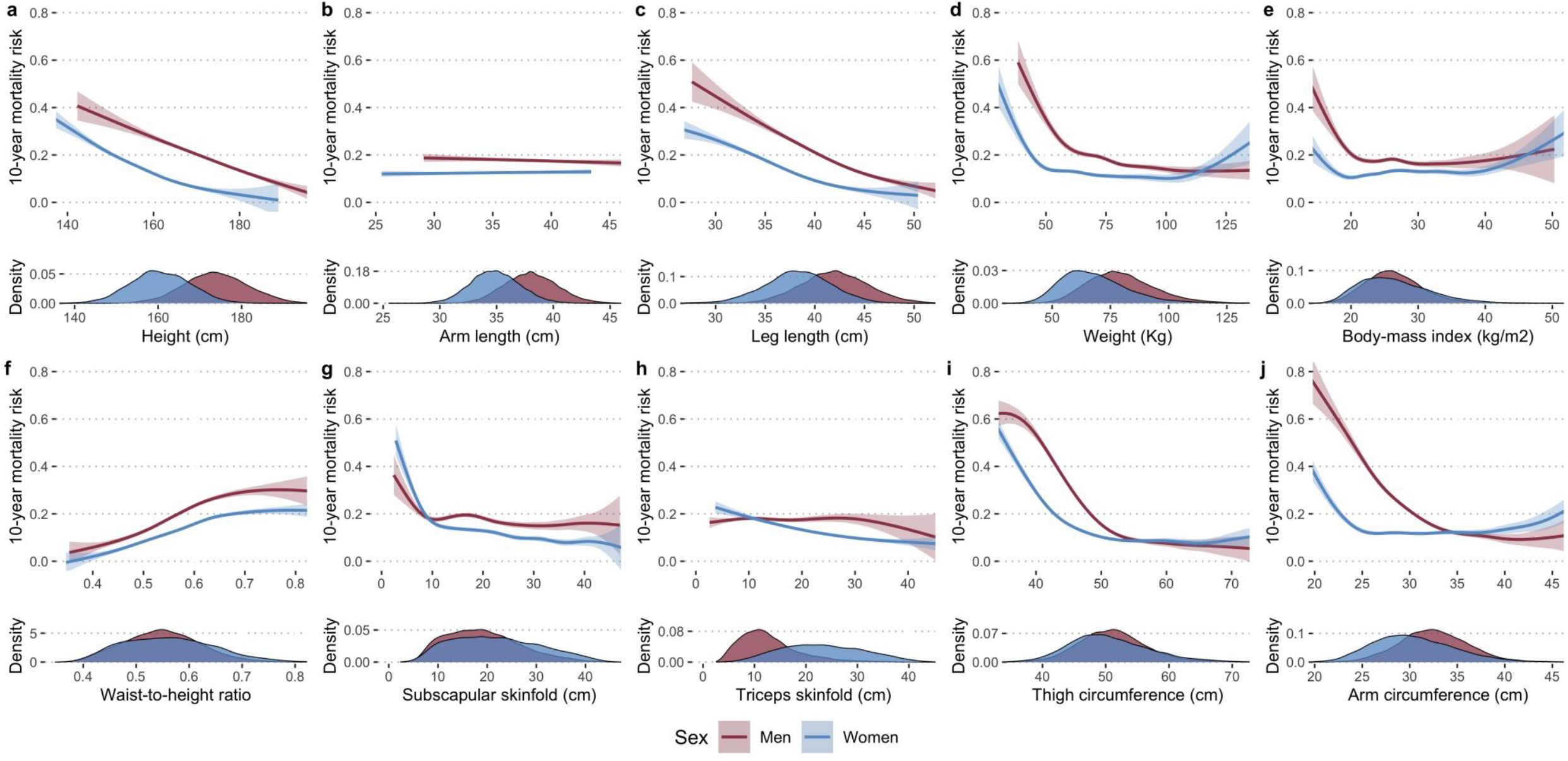
Anthropometric variables predict 10-year all-cause mortality in a non-linear and sex-dependent fashion. Variables are grouped according to the property of body composition that they capture: body length (a-c), body mass (d-e), visceral adiposity (f), subcutaneous adiposity (g-h), primarily lean mass (i-j). We used Gompertz models stratified by sex and adjusted by CA and number of comorbidities, with race/ethnicity included in the shape parameter.

### Anthropometric measurements are associated with sex-specific mortality patterns

We first sought to comprehensively characterize the non-linear relationship between anthropometric measurements and mortality risk independently of CA, comorbidities and race/ethnicity using Gompertz proportional hazards models (see **Methods**). Body lengths, particularly height and leg length, displayed an inverse linear relationship with mortality (**Figure 2a-c**). Weight and BMI had similar U-shaped curves as previously reported in numerous studies (Donini et al., 2020; Lavie et al., 2009), where mortality risk was the highest at lower values and virtually invariant thereafter, except for a small increase at extremely high values (particularly in women) (**Figure 2d-e**). Visceral adiposity measured by WHtR was associated with a progressive risk increase for both genders (**Figure 2f**), while higher reserves of subcutaneous adiposity were related to a marked decline in mortality risk in women, and to a lesser extent in men (**Figure 2g-h**). Interestingly, mortality showed a similar behavior with arm and thigh circumferences as the one seen for BMI (**Figure 2i-j**); we initially established that these variables would capture primarily lean mass, but this has been shown to be influenced by age, gender, and total adiposity (Cavedon et al., 2020). Thus, lower values of BMI, arm and thigh circumferences confer a dramatic increase in mortality likely due to reduced muscle mass, while the slight risk increase observed at greater values could be attributed to higher fat-mass. In summary, highest mortality was observed for decreased total and lean mass, moreover, in most cases men had a higher mortality risk than women except for instances of very low subcutaneous adiposity and extreme obesity.

### Estimation of Anthropometric Age

We then developed our own BA estimator which we termed Anthropometric Age (AnthropoAge) by adapting the methods previously described to develop PhenoAge using Gompertz models (Levine et al., 2018). As the relationship between anthropometry and mortality risk is heavily influenced by sex, we decided to perform variable selection separately for men and women. In women, AnthropoAge includes weight, WHtR, thigh circumference, subscapular skinfold, and triceps skinfold; while in men it only includes WHtR, thigh circumference, and arm circumference (**Supplementary Table 3**). Our data driven approach confirms that anthropometric variables contribute to BA and mortality risk in a sex-dependent fashion. Roughly, the best predictors are variables that capture visceral adiposity and lean mass for men, and variables that capture visceral and subcutaneous adiposity for women. Anthropometry is an unexpensive and easily implementable tool for clinical practice (Padilla et al., 2021); however, some of these measurements are not as commonly used. To promote a wider application of the concept of anthropometric aging into large-scale epidemiological studies, we sought to develop a simpler version of AnthropoAge using exclusively BMI and WHtR given the robust association of changes in body fat distribution with aging (Bello-Chavolla et al., 2020; Tchernof & Després, 2013) and the relationships that these metrics displayed with mortality risk; the resulting metric was termed simplified AnthropoAge (S-AnthropoAge) (**Supplementary Table 4**). AnthropoAge and S-AnthropoAge act as a proxy of BA and represent the predicted 10-year mortality risk based on an individual’s anthropometry. Notably, we found no significant discrepancies or systematic bias between AnthropoAge and S-AnthropoAge, with overall low bias (0.239 years, 95% CI: 0.208 to 0.270) and a high intra-class correlation coefficient (ICC 0.9940, 95%CI 0.9937-0.9943, p<0.001) (**Supplementary Figure 3**).

**FIGURE 3.**
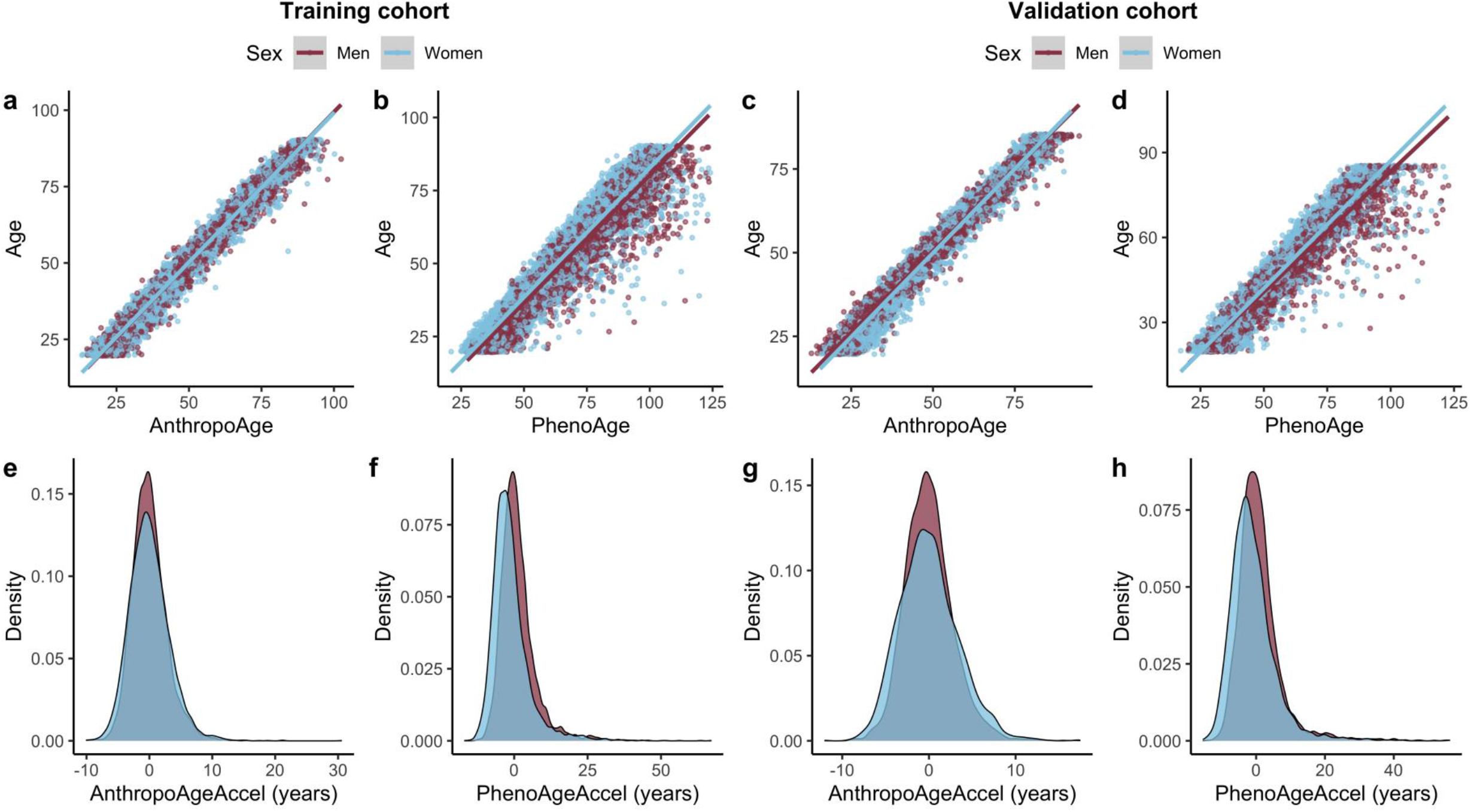
AnthropoAge and PhenoAge scatter plots show that these BA metrics have different dispersion patterns with CA in both the training (a-b) and validation cohorts (c-d). The horizontal distances between each point and regression lines represent divergence from CA and were used to estimate age acceleration metrics, which are presented as density plots stratified by sex (e-h). Only AnthropoAgeAccel was calculated separately for each sex, as reflected by a more homogeneous distribution between men and women.

### Estimation of Anthropometric Age Acceleration

Next, we calculated Phenotypic Age Acceleration (PhenoAgeAccel) as described elsewhere (Liu et al., 2018) and followed this method to obtain a new metric of age acceleration that we called Anthropometric Age Acceleration (AnthropoAgeAccel and S-AnthropoAgeAccel for the simplified version). These metrics represent the divergence of BA from CA, where values >0 indicate a 10-year mortality risk greater than that predicted by CA (accelerated aging) and values ≤0 represent an equal or lower risk (physiological aging). All of our newly developed metrics (AnthropoAge, S-AnthropoAge, AnthropoAgeAccel and S-AnthropoAgeAccel) have been deployed within a ShinyApp along with the estimation of PhenoAge to facilitate their use for clinical and research purposes, available at https://bellolab.shinyapps.io/anthropoage/.

### BA estimates across sex and race/ethnicities

There were no significant differences between men and women in AnthropoAge (45.3 years [IQR: 31.7 – 62.6] vs. 44.4 years [IQR: 31.5 – 62.8], p=0.671) nor AnthropoAgeAccel (−0.18 years [IQR: -1.88, 1.57] vs -0.21 years [IQR: -2.31, 2.04], p=0.4406) in the validation cohort. In contrast, both PhenoAge (56.3 years [IQR: 41.7 – 74.1] vs. 53.7 years [IQR: 39.4 – 71.2], p<0.001) and PhenoAgeAccel (−0.08 years [IQR: -2.92, 3.13] vs -2.01 years [IQR: -5.19, 1.79], p<0.001) were significantly increased in men (**Figure 3**). On the other hand, we compared age acceleration metrics across race/ethnicities, and observed higher PhenoAgeAccel values in non-Hispanic Black participants, while AnthropoAgeAccel and S-AnthropoAgeAccel were higher among Mexican Americans (**Supplementary Figure 4**).

**FIGURE 4.**
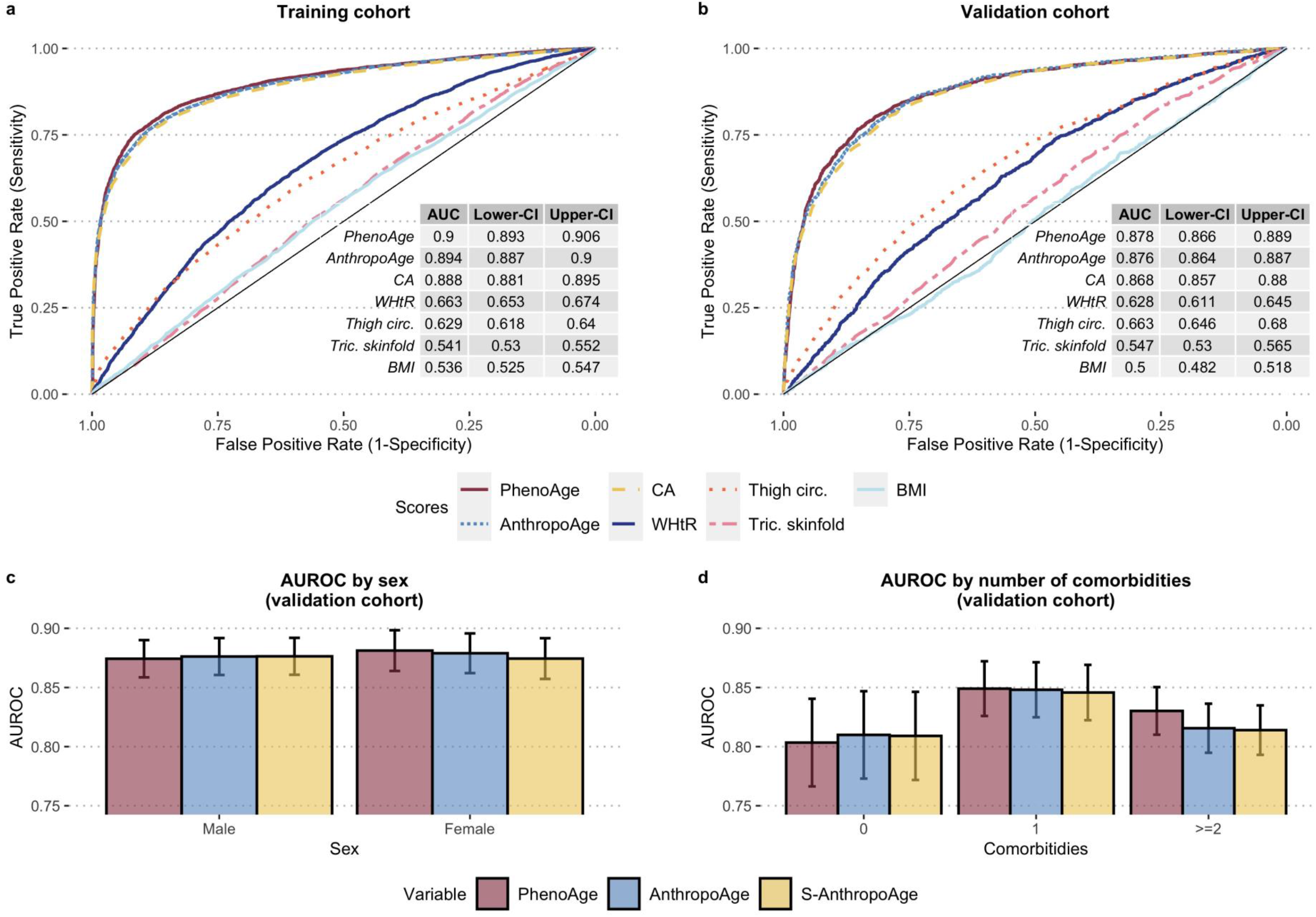
Areas under the receiving operating characteristic curves (AUROC) for prediction of 10-year all-cause mortality demonstrate that AnthropoAge and S-AnthropoAge (not shown) have a comparable performance to PhenoAge, and that it outperforms CA and individual anthropometric measurements (WHtR, thigh circumference, triceps skinfold and BMI) (a-b). This prediction is equally good between men and women (c), but superior in subjects with 1 comorbidity (b). There were no significant differences between the three BA metrics in any category except for subjects with ≥2 comorbidities.

### AnthropoAge and S-AnthropoAge predict 10-year all-cause mortality

We evaluated the performance of AnthropoAge for prediction of all-cause mortality compared to PhenoAge, CA and individual anthropometric measurements. AnthropoAge had a significantly higher AUROC compared to CA, BMI, WHtR, thigh circumference and triceps skinfold. In the training cohort, AnthropoAge had a significantly lower AUROC compared to PhenoAge (p<0.001), however, they had a similar performance in the validation cohort (p=0.307) **(Figure 4a-b)**. When assessing AUROC stratified by sex and by number of comorbidities, we observed that AnthropoAge performed similarly between men and women and had a better performance for subjects with 1 comorbidity. PhenoAge was slightly superior to AnthropoAge in subjects with ≥2 comorbidities (p=0.041), but there were no significant differences in any other subgroup (**Figure 4c-d, Supplementary Table 5**).

### Cause-specific mortality is heterogeneously predicted by different aging metrics

We hypothesized that AnthropoAge and PhenoAge would assess different domains of aging. For this purpose, we evaluated whether AnthropoAge and PhenoAge were better at predicting distinct mortality causes. All metrics were overall strong to predict cause-specific mortality after adjustment by sex, ethnicity, and number of chronic comorbidities. However, AnthropoAge and S-AnthropoAge had better predictive performance for cerebrovascular, Alzheimer’s disease and chronic lower respiratory disease related mortality, while PhenoAge was superior for diabetes and nephritis/nephrosis related mortality. All metrics had a roughly comparable performance for cardiovascular, cancer and influenza/pneumonia related mortality (**Table 1, Supplementary Table 6**). Next, we evaluated a possible interaction effect between BA and sex on cause-specific mortality. We found that AnthropoAge significantly interacted with female sex to predict cardiovascular disease (HR for interaction 1.014, 95%CI: 1.007-1.020), diabetes mellitus (HR for interaction 1.028, 95%CI: 1.014-1.042), stroke (HR for interaction 1.022, 95%CI: 1.010-1.034), and cancer (HR for interaction 0.989, 95%CI: 0.984-0.994) related mortality. These results a) suggest that AnthropoAge and PhenoAge could be assessing different aspects of aging and b) strengthen the notion that sex dimorphisms link changes in body composition to poor age-related outcomes.

### Multidomain age acceleration increases mortality risk and comorbidity burden

We also explored mortality trajectories of subjects with and without accelerated aging using Kaplan-Meier curves. Notably, subjects with accelerated aging displayed higher risk of overall mortality compared to those with physiological aging; this result was replicated for all age acceleration metrics (**Figure 5a-c**). Furthermore, we built an indicator of multidomain age acceleration with the following categories: physiological aging (both AnthropoAgeAccel and PhenoAgeAccel ≤0), accelerated AnthropoAge (AnthropoAgeAccel >0 but PhenoAgeAccel ≤0), accelerated PhenoAge (AnthropoAgeAccel ≤0 but PhenoAgeAccel >0) and multidomain acceleration (both AnthropoAgeAccel and PhenoAgeAccel >0) and found that individuals with multidomain acceleration had the highest risk of mortality (**Figure 5d**). Cases with accelerated AnthropoAge (HR 1.46, 95%CI: 1.35-1.59), accelerated PhenoAge (HR 1.74, 95%CI: 1.60-1.88) and multidomain acceleration (HR 2.43, 95%CI: 2.25-2.62) had higher risk for all-cause mortality compared to individuals with physiological aging after adjustment for age, sex, number of comorbidities, and ethnicity. A similar pattern was observed with S-AnthropoAge (HR for multidomain acceleration: 2.22, 95%CI: 2.06-2.40). Aging metrics have been previously shown to be associated with comorbidity profiles (C. Kuo et al., 2021; Liu et al., 2018; Sayed et al., 2021), on that basis, we investigated whether specific comorbidity profiles were captured by anthropometric aging. Subjects with an increasing number of comorbidities had higher AnthropoAgeAccel and S-AnthropoAgeAccel independently of CA categories (**Supplementary Figures 5a, 6a**). Similarly, when using the multidomain aging indicator, subjects had a higher comorbidity burden in the multidomain acceleration category, indicating that simultaneous consideration of these metrics increases the likelihood of identifying accumulation of comorbidities (**Supplementary Figures 5b, 6b**).

**FIGURE 5.**
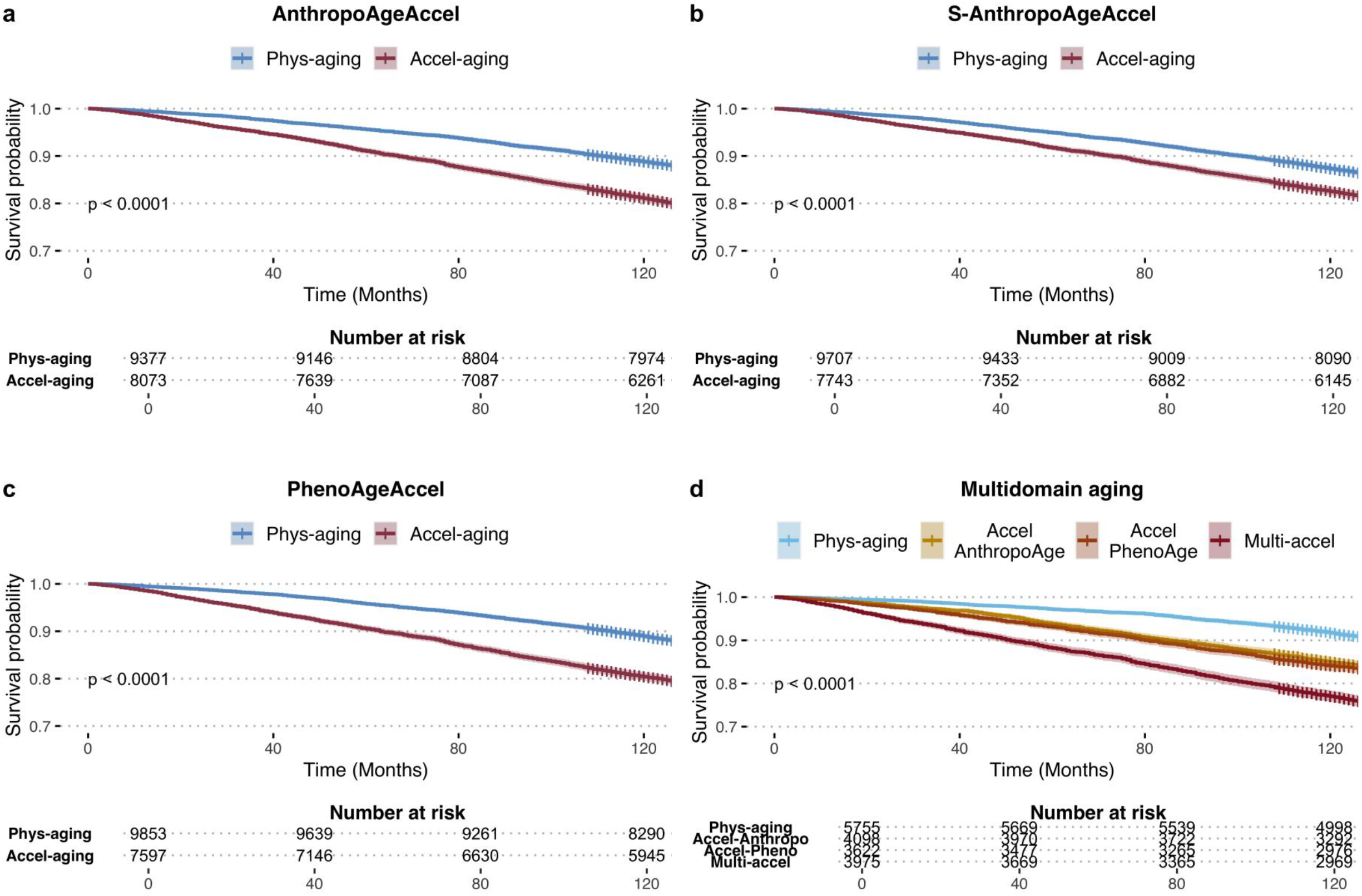
AnthropoAgeAccel (a), S-AnthropoAgeAccel (b) and PhenoAgeAccel (c) identify subjects with impaired survival linked to accelerated aging (values >0) visualized with Kaplan-Meier curves. Simultaneous consideration of AnthropoAgeAccel and PhenoAgeAccel shows that acceleration in multiple domains of aging produces a higher mortality risk (d).

### Accelerated aging determines specific body composition and biochemical phenotypes

Next, we characterized body composition and biochemical profiles in 6,284 cases with complete DXA information (**Figure 1**). We stratified individuals by sex and compared those with AnthropoAgeAccel >0 (accelerated anthropometric aging) to those with AnthropoAgeAccel ≤0. These variables are presented as spider plots to facilitate interpretation and pattern-recognition (**Figure 6**), the following description summarizes the most relevant changes found in subjects with accelerated aging. In women, accelerated anthropometric aging was related to a larger overall body mass (comprising fat and lean mass), and an important accumulation of visceral adiposity (greater fat mass and fat-to-lean ratio in trunk, and greater WHtR) disproportionate to their subcutaneous fat reserves (smaller skinfolds, greater trunk-to-appendicular fat mass ratio); notably, these women also presented larger amounts of total and appendicular lean mass. Men with accelerated aging had markedly lesser amounts of fat and lean mass in all body compartments (the most dramatic difference was seen in appendicular lean mass). Although they had a lower subcutaneous adiposity, evidence of visceral fat accumulation was negligible; they also presented reduced bone mineral density. On the other hand, laboratory data showed that women with accelerated anthropometric aging had a distinctive proinflammatory profile with greater CRP, WBC and ALP and lower albumin and lymphocyte percentage; they also had higher PhenoAge values. Meanwhile, in men with accelerated anthropometric aging the aforementioned changes were not present or were modest (ALP, WBC), however, these subjects had a significantly lower creatinine, which could be attributed to less muscle mass. Using S-AnthropoAgeAccel led to similar body composition patterns linked to accelerated aging, with the only major changes being 1) a more prominent accumulation of abdominal fat and reduction of bone mineral density in both sexes, and 2) no increase in lean mass in women with accelerated aging (**Supplementary Figure 7**). Finally, we sought to evaluate how multidomain acceleration would impact on body composition and biochemical profiles and found that most of the changes described above seem to be greatly augmented when both AnthropoAge and PhenoAge are accelerated (**Supplementary Figure 8**). Our findings denote sharp sexual dimorphisms in body composition and inflammatory biomarkers in individuals at higher risk of mortality, which requires further evaluation.

**FIGURE 6.**
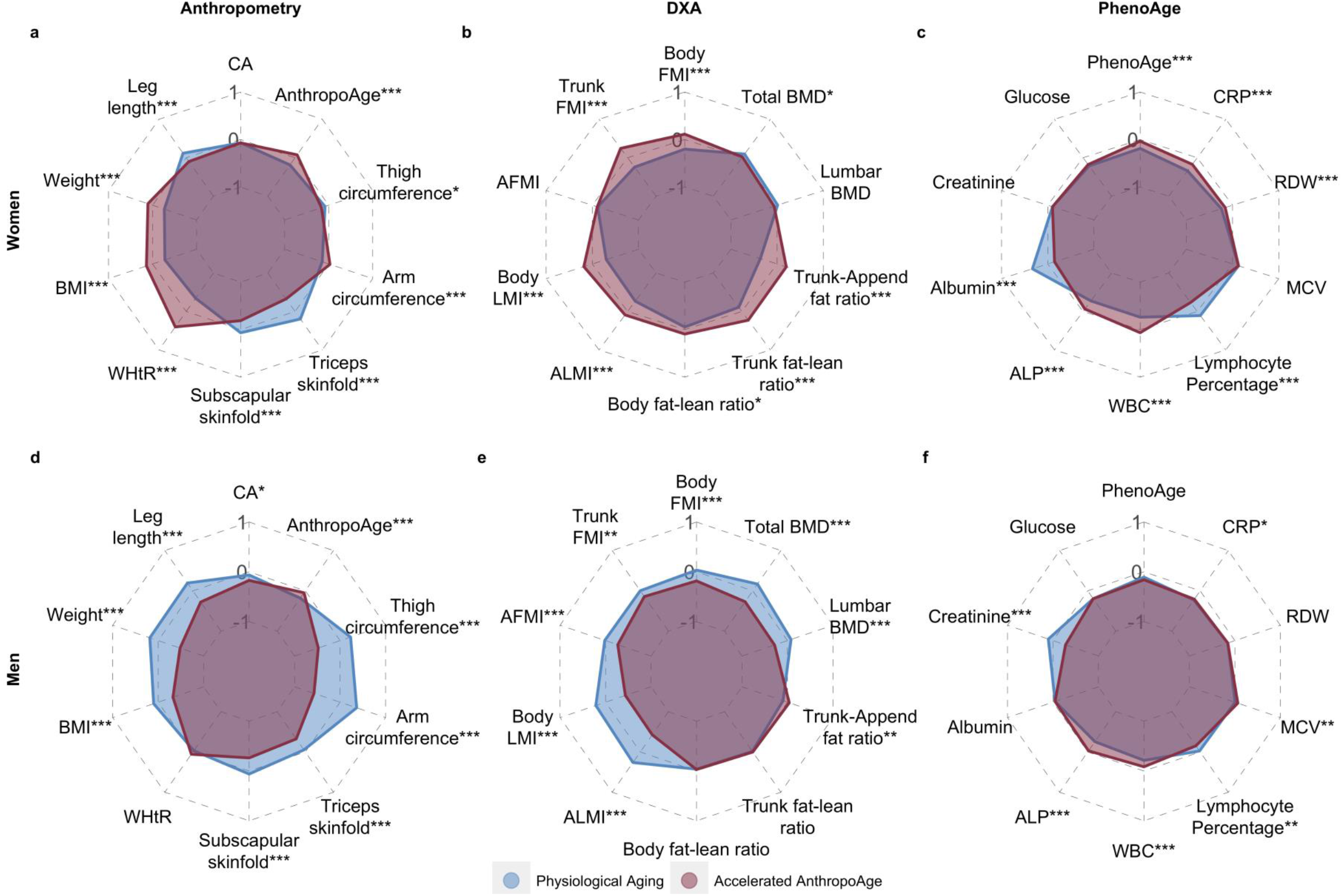
Spider plots stratified by sex show that subjects with accelerated AnthropoAge (AnthropoAgeAccel >0) display unique patterns of anthropometry, DXA-derived body composition and biochemical profiles (PhenoAge components). DXA variables were divided by squared height to obtain whole-body, trunk and appendicular fat mass and lean mass index (FMI and LMI, respectively), we also calculated fat-to-lean mass ratios and trunk-to-appendicular fat ratio. Variables were scaled and medians were compared using Mann-Whitney’s U test (p-value <0.05: *, <0.01: **, <0.001: ***). Spider plots assessing S-AnthropoAgeAccel and multidomain aging are shown in supplementary figures 7 and 8.

## DISCUSSION

In this work, we developed a novel aging metric based on anthropometric parameters, aiming to predict 10-year mortality risk as a proxy of BA. We showed that AnthropoAge and S-AnthropoAge predict all-cause mortality independently of sex, CA, ethnicity, and number of chronic comorbidities. Notably, AnthropoAge and S-AnthropoAge were better predictors for Alzheimer’s, stroke, and chronic lower respiratory related mortality than PhenoAge. We also found that subjects with accelerated anthropometric aging have sharp sexual dimorphisms in body composition and in cause-specific mortality risk. Finally, we demonstrate that PhenoAgeAccel and AnthropoAgeAccel, when considered simultaneously, may identify aging phenotypes with unique comorbidity profiles and differential risk of all-cause mortality, which may indicate domain or tissue-specific aging rates (Ahadi et al., 2020; C. Kuo et al., 2021).

Body composition has been previously explored for the evaluation of BA, identifying sex-based differences of anthropometric parameters that are linearly associated with CA (Kang et al., 2012; Negasheva et al., 2017). In contrast to these previous studies, AnthropoAge uses a non-linear approach to link body composition patterns to mortality risk, which has been speculated to be a better surrogate of BA; this in turn may clarify the relationship of body composition with aging and age-related outcomes beyond individual metrics such as BMI, given that these alone do not completely capture the complexity of this phenomenon (Donini et al., 2020; Lavie et al., 2009). Thus, inclusion of a richer diversity of variables paired with refined modeling methods may lead to more precise assessments of BA (Ahadi et al., 2020; Elliott et al., 2021; Sayed et al., 2021; Xia et al., 2020).

In addition to aging mechanisms that may lead to changes in body composition, numerous studies have also drawn connections between body features and poor age-related outcomes such as disability, age-related diseases, and mortality (Kim & Won, 2022; Santanasto et al., 2016; Schorr et al., 2018). Evidence suggests that the bidirectional relationship between aging and body composition can eventually turn into a vicious cycle that results in an aggravation of age acceleration (Salvestrini et al., 2019), and that this phenomenon is heavily influenced by sex dimorphisms (Goossens et al., 2021; MAGIC et al., 2010; Pomatto et al., 2018; Sampathkumar et al., 2020; Wells, 2007). A relevant addition of our study is that we characterized sexual dimorphisms in the context of accelerated anthropometric aging. Particularly, we observed that females with accelerated aging display increased body mass and a disproportionately larger collection of visceral fat in relation to subcutaneous fat, while males present a phenotype of deeply decreased lean and fat mass with a mild accumulation of visceral adiposity. Furthermore, anthropometric aging significantly increased cardiovascular, diabetes and stroke-related mortality for women, and cancer-related mortality for men. A plausible explanation for these findings could be post-menopausal estrogens loss, which have been shown to elicit beneficial effects on several processes such as glucose and lipid metabolism in skeletal muscle, adipose tissue, and liver (Goossens et al., 2021), bone homeostasis (Khosla & Monroe, 2018), and cognitive function (Gurvich et al., 2018), among others. Peripheral estrogen production in subcutaneous adipose tissue can offer some of these beneficial effects (Sampathkumar et al., 2020), as a result, a marked shift from subcutaneous to visceral adiposity could precipitate cardiometabolic risk (Goossens et al., 2021; Schorr et al., 2018), as seen in women with accelerated aging from our study. On the other hand, men have been reported to have less muscle mitochondrial content and activity (Rosa-Caldwell & Greene, 2019) and greater systemic proinflammatory responses (Márquez et al., 2020), which may increase their susceptibility for inflammation-induced muscle loss as seen in several age-related diseases including chronic obstructive pulmonary disease and cancer (Machado et al., 2021; Montalvo et al., 2018). Thus, AnthropoAge, S-AnthropoAge and their age acceleration metrics widen our understanding of disparities in aging rates and longevity attributable to sex. Further studies are required to determine the extent to which sex differences in fat distribution and muscle functionality may aid in the estimation of BA and its relative performance in comparison to blood biomarkers.

A previous analysis using data from the Baltimore Longitudinal Study of Aging proposed four domains to integrate functional, phenotypic, and biological aging rates including: body composition, energy regulation, homeostatic mechanisms, and neurodegeneration/neuroplasticity (P. -L. Kuo et al., 2020). Similarly, the work by Kuo et al. demonstrated that PhenoAge and BioAge evaluate different domains of aging, with genetic data suggesting unique aging pathways for each BA indicator (C. Kuo et al., 2021; Levine, 2013). In line with these findings, we observed that PhenoAge and AnthropoAge have distinct abilities to predict cause-specific mortality; furthermore, simultaneous consideration of both accelerated metrics establishes unique mortality risk trajectories and comorbidity profiles, which may better reflect the heterogeneity of aging rates. Consideration of multiple markers of aging may increase the likelihood of modeling complex interactions between different biological domains and it may prove useful to tailor specific strategies to reduce the burden associated with unhealthy aging (Scott et al., 2021). Our newly developed metric could represent a step further in this direction by providing a precise and easily implementable estimation of the body composition domain of biological aging.

This study has numerous strengths that support the validity and clinical relevance of our findings. By using large population-based data, we were able to capture a diverse population, with a wide-range of body phenotypes and varying comorbidity burden. Furthermore, we aimed to shed light on the multidimensionality of the aging process by contrasting and combining multiple aging metrics. Lastly, AnthropoAge and S-AnthropoAge offer unique opportunities to translate the concept of BA onto clinical practice or routine use, as they can be implemented with simpler measurements compared to those using omics technologies or genomic data (Jansen et al., 2021; Li et al., 2020; Rivero-Segura et al., 2020). Nonetheless, our results are subject to some limitations. First, despite taking races/ethnicities into account to estimate AnthropoAge and S-AnthropoAge, we found substantial differences in aging rates across racial and ethnic categories, this implies that specific validation studies may be required to adapt these metrics to different populations due to the marked racial and ethnic diversity in aging rates and body composition (Caleyachetty et al., 2021). Second, we were not able to assess how longitudinal changes in AnthropoAge, S-AnthropoAge and their accelerated metrics modify mortality risk; future longitudinal studies may allow to translate these metrics onto specific strategies to intervene on aging rates by targeting body composition. Third, despite being readily accessible, the reliability of anthropometric parameters depends upon adequate techniques and reproducibility for their measurement, which may hinder their adequate usage (Wang et al., 2000).

In conclusion, we characterized the CA-independent contribution of body composition to mortality risk for men and women and implemented anthropometric measurements into the estimation of biological age, resulting in the development of complete and simplified versions of AnthropoAge and its age acceleration indicator AnthropoAgeAccel. These BA estimators capture mortality risk, comorbidity profiles and identify body composition phenotypes with marked sexual dimorphisms linked to accelerated aging, and they can be easily implemented into clinical practice to monitor and promote healthier aging. Finally, we showed that consideration of multiple BA metrics may address the aging process from different perspectives, thus improving the identification of aging trajectories and shedding light on its complexity and heterogeneity.

## METHODS

### NHANES

NHANES is a nationally representative cross-sectional survey designed to assess health and nutritional status of US population conducted in waves by the Centers for Disease Control and Prevention (CDC). NHANES-III refers to the third wave conducted from 1988 to 1994, and NHANES-IV (also known as continuous NHANES) corresponds to the ongoing wave, which comprises 2-year cycles starting in 1999 (in this study we used data up to 2008). NHANES underwent National Center for Health Statistics (NCHS) Research Ethics Review Board approval, and all participants provided informed consent. Complete methods for recruitment, procedures, and study design for NHANES are described in detail elsewhere (*NHANES - National Health and Nutrition Examination Survey Homepage*, 2022). This project was registered and approved by the Ethics and Research Committee at Instituto Nacional de Geriatría, project number DI-PI-006/2020.

### Body composition data

In addition to demographic, socioeconomic and health-related questionnaires, a subset of NHANES underwent further biochemical and anthropometric evaluations. We only included anthropometric measurements available in both NHANES-III and IV (since calf and hip circumference were not available in all cycles, these parameters were not considered for this study). Normal distribution was assessed with the Anderson-Darling test and variable transformations were performed as specified in **Supplementary Figure 1**; an overview of measuring techniques is provided in **Supplementary Table 1**. For a subset of NHANES-IV, whole-body Dual X-ray Absorptiometry (DXA) assessments were acquired using Hologic QDR 4500A fan-beam bone densitometers.

### All-cause and cause-specific mortality data

We obtained all-cause mortality follow-up data using information from the National Death Index for NHANES-III and IV through March 2020. Cause-specific mortality information was evaluated for 8 out of 10 underlying causes of death: cardiovascular, chronic lower respiratory disease, cerebrovascular disease, malignant neoplasms, Alzheimer’s disease, diabetes mellitus, influenza/pneumonia, and nephritis/nephrosis; excluding accidents and non-specified causes of death (*NCHS Data Linkage - Mortality Data*, 2022). Follow-up time was estimated from date of initial interview to last follow-up in person-month time.

### Estimation of biological age

#### PhenoAge

PhenoAge was previously developed in NHANES-III and validated in NHANES-IV by Levine et al. (Levine et al., 2018; Liu et al., 2018) using a Gompertz proportional hazards model to predict 10-year mortality risk based on CA and the nine blood biomarkers previously mentioned. We obtained PhenoAge values by using the equation described elsewhere (Liu et al., 2019).

#### AnthropoAge

We used a similar approach to develop AnthropoAge using proportional hazards regression models with the parametric Gompertz distribution. First, we fitted two Gompertz models to predict all-cause 10-year mortality risk: the first one had only CA as predictor, and the second had anthropometric measurements as predictors. We equated the risk from both models and solved for age to convert the mortality risk into units of years, thus obtaining AnthropoAge (for a detailed description see **Supplementary Methods**). Importantly, we used orthogonal polynomials to model the non-linear relationship between anthropometry and mortality risk, adjusted for CA and with ethnicity being included in the shape parameter of the Gompertz distribution. To select which anthropometric variables would be included and the optimal number of degrees for orthogonal polynomials, we systematically tested all possible combinations and chose the best model according to minimization of the Bayesian Information Criterion (BIC). We controlled for variable correlation (**Supplementary Figure 2**) and multicollinearity by dropping variables with a Variance Inflation Factor (VIF) ≥10. All Gompertz models were carried out with the *flexsurv* R package, which allows modeling of time-to-event data using any parametric distribution; in particular, the Gompertz distribution assumes an exponential rise in mortality rate as the population ages (Jackson, 2016; Wilson, 1994). To assess average bias and limits of agreement between AnthropoAge and S-AnthropoAge we carried out Bland-Altman analyses with the *blandr* R package.

### Estimation of age acceleration

Following the methods by Liu et al. used to calculate PhenoAgeAccel (Liu et al., 2018), we developed a metric to estimate aging acceleration. We regressed AnthropoAge onto CA in a least-squares linear regression and extracted residuals from this model, which represent the deviation of anthropometric age from CA; a noteworthy difference from method originally described is that AnthropoAgeAccel and S-AnthropoAgeAccel were calculated separately for men and women.

### Assessment of AnthropoAge performance

We validated the utility of AnthropoAge for prediction of 10-year all-cause mortality in NHANES-IV using Gompertz proportional hazard regression models and the area under receiver operating characteristic curves (AUROC) and compared its performance with PhenoAge, CA, and individual anthropometric measurements using non-parametric ROC tests with bootstrapping (B=1,000) in the *pROC* R package (Robin et al., 2011).

### Cause-specific mortality analysis

We evaluated cause-specific mortality using Fine & Gray semiparametric competitive risk regression models with the *survival* R package (Therneau et al., 2022) to contrast predictive performance between AnthropoAge, S-AnthropoAge and PhenoAge in subjects with complete mortality data. All models were adjusted by sex, ethnicity, and number of chronic comorbidities. Predictive performance was assessed by the C-statistic and BIC differences (ΔBIC = BIC_Model-1_ - BIC_Model-2_), where a ΔBIC < -2 indicates that the first model is better, a ΔBIC > 2 indicates that the second model is better, and a ΔBIC in between indicates that both models are comparable for the prediction of a specific outcome.

### Assessment of mortality trajectories

We used Kaplan-Meier curves and the log-rank test with the *survival* R package to compare mortality trajectories between subjects with physiological and accelerated aging for all metrics (AnthropoAgeAccel, S-AnthropoAgeAccel and PhenoAgeAccel). We also plotted Kaplan-Meier curves to compare mortality between categories from the multidomain aging indicator.

### Body composition and biochemical phenotypes

We evaluated sex-specific differences in body composition and biochemical profile between subjects with accelerated and non-accelerated aging using laboratory data, anthropometry and DXA measurements from NHANES-IV cycles. Regarding DXA, we divided fat mass and lean mass variables by squared height, resulting in standardized fat mass and lean mass indexes (FMI and LMI, respectively); this was done for whole-body, trunk and appendicular compartments. We also calculated fat-to-lean ratios (fat mass divided by lean mass) in whole-body and trunk and a trunk-to-appendicular fat ratio (trunk fat mass divided by appendicular fat mass) to assess disproportionate fat accumulations. All variables were Z-transformed to facilitate visualization using spider plots with the *fmsb* R package.

Comparisons between categorical and continuous variables were performed with the chi-squared and the Mann-Whitney U test, respectively. All statistical analyses were conducted using R version 4.1.1 and p-values thresholds are estimated for a two-sided significance level of α=0.05.

## Data Availability

All code, datasets and materials are available for reproducibility of results at https://github.com/oyaxbell/anthropoage/

## ACKNOWLEDGMENTS

This project was registered and approved by the Research Committee at Instituto Nacional de Geriatría, project number DI-PI-006/2020. NEAV, CAFM, AMS, ECG, and ESC are enrolled at the PECEM Program of the Faculty of Medicine at UNAM. NEAV and ESC supported by CONACyT. The graphical abstract and figure 1 were designed using resources created by *Victoruler, Smashicons, Freepik* and *Surang* from www.flaticon.com

## AUTHOR CONTRIBUTIONS

Research idea and study design: CAFM, AMS, ECG, NEAV, LMGR, OYBC; data acquisition: CAFM, AMS, ECG, OYBC; analysis/interpretation: CAFM, AMS, OYBC; statistical analysis: CAFM, AMS, OYBC; manuscript drafting: CAFM, AMS, ECG, LZR, LFC, DABG, NEAV, ESC, ACM, AVV, CDPC, DRG, LMGR, OYBC; supervision or mentorship: OYBC. Each author contributed important intellectual content during manuscript drafting or revision and accepts accountability for the overall work by ensuring that questions pertaining to the accuracy or integrity of any portion of the work are appropriately investigated and resolved.

