## Supplementary Material for "AnthropoAge, a novel approach to integrate body composition into the estimation of biological age"

### SUPPLEMENTARY METHODS

#### Derivation of AnthropoAge and Simplified AnthropoAge (S-AnthropoAge)

*The following derivations are based upon the methods proposed by Levine et al. Given the well-known differences in body composition across sexes and the influence of ethnicity on body composition traits, we fitted Gompertz proportional hazard regression models stratified by sex and with ethnicity in the sape parameter. We used the flexsurv R package to estimate rate, shape, and β-coefficients for both AnthropoAge and S-AnthropoAge. The procedure to extract the formula is as follows:*

1) *Gompertz CDF (cumulative distribution function) for 10-year mortality risk using chronological age as predictor:*

$$\mathrm{CDF}(120,\mathrm{age}) = 1 - \mathrm{e}^{-\left(\mathrm{e}^{\mathrm{age}*\beta_1+\beta^0}\gamma_0{}^{-1}\left(\mathrm{e}^{\gamma_0 t}-1\right)\right)}$$

2) *Gompertz CDF for 10-year mortality risk using anthropometric scores (xb) as predictors:*

$$\mathrm{CDF}(120,\mathrm{xb}) = 1 - \mathrm{e}^{-\left(\mathrm{e}^{\mathrm{xb}}\gamma_1{}^{-1}\left(\mathrm{e}^{\gamma_1 t}-1\right)\right)}$$

3) *To derive Anthropometric Age (AnthropoAge) we equate the Gompertz CDF for chronological age to the Gompertz CFD for anthropometric score at 120 months (10 years).*

$$\mathrm{CDF}(120,\mathrm{age}) = \mathrm{CDF}(120,\mathrm{xb})$$

4) *The Gompertz function assumes an exponential growth in mortality as a population ages, and its CDF can be written as:*

$$1 - \mathrm{e}^{-\mathrm{e}^{\mathrm{age}*\beta_1+\beta^0}\gamma_0{}^{-1}\left(\mathrm{e}^{\gamma_0 t}-1\right)} = \mathrm{CDF}(120,\mathrm{xb})$$

5) *Now, we let* $M = CDF(120, xb)$

$$1 - \mathrm{e}^{-\left(\mathrm{e}^{\mathrm{age}*\beta_1+\beta^0}\gamma_0{}^{-1}\left(\mathrm{e}^{\gamma_0 t}-1\right)\right)} = \mathrm{M}$$

6) *We take the natural logarithm of both sides and multiply by -1*

$$-e^{age*\beta_1+\beta^0}{\gamma_0}^{-1}(e^{\gamma_0 t}-1)=\ln(1-M)$$

7) *Next, we isolate the term $e^{age*\beta_1+\beta^0}$ on the left-hand side*

$$-e^{age*\beta_1+\beta^0}=\frac{\ln(1-M)}{{\gamma_0}^{-1}(e^{\gamma_0 t}-1)}$$

8) *We then take the natural logarithm of both sides of the equation*

$$age*\beta_1+\beta^0=\ln\left(-\frac{\ln(1-M)}{{\gamma_0}^{-1}(e^{\gamma_0 t}-1)}\right)$$

9) *We isolate age and define AnthropoAge as follows:*

$$\text{AnthropoAge}=\frac{\ln\left(-\frac{\ln(1-M)}{{\gamma_0}^{-1}(e^{\gamma_0 t}-1)}\right)-\beta^0}{\beta_1}$$

**Variable transformation**

To improve model diagnostics, we conducted variable transformations for anthropometric variables by evaluating logarithmic, squared root, cubic root, inverse, and exponential transformations. Optimal transformations were chosen by minimization of the A-statistic from the Anderson-Darling normality test. The following anthropometric measurements were transformed using natural logarithms: weight, body mass index, arm length and thigh circumference. Square root transformation was used for arm circumference and cubic root transformation was used for height, triceps and subscapular skinfolds, and waist-to-height ratio. Leg length did not require any specific transformation (**Supplementary Figure 1**). All models were fitted using transformed variables unless indicated otherwise. Orthogonal polynomials were fitted to introduce non-linearity to the models using the *poly* R function and the number of degrees of freedom was arrived at by using BIC minimization as the model selection criterion.

### SUPPLEMENTARY TABLES

**SUPPLEMENTARY TABLE 1.** Brief description of the measuring techniques for all anthropometric variables included in our study. Variables are grouped according to the body composition property they capture. Detailed descriptions are available at the NHANES anthropometry procedures manual: https://wwwn.cdc.gov/nchs/data/nhanes/1999-2000/manuals/bm.pdf

| Body composition property | Anthropometric measurement | Measuring technique |
|---|---|---|
| **Body length** | Height (cm) | The subject is positioned standing with both feet flat on the floor, heels together and toes pointed slightly outwards, looking straight forward and with head, shoulder blades, buttocks, and heels touching the measurement surface. The head is aligned until horizontal line from the ear canal to the lower border of the orbit of the eye is parallel to the floor. Height is recorded to the nearest millimeter with a fixed stadimeter (headboard positioned on top of the head with enough pressure to compress the hair). |
| | Upper arm length (cm) | With the subject standing (right arm flexed at 90 degrees and palm facing up), the length is recorded to the nearest millimeter from the posterior border of the acromion process of the right scapula to the tip of the right olecranon process. A mark is placed at the midpoint to define where arm circumference and tricipital skinfold will be measured. |
| | Upper leg length (cm) | With the subject sitting (lower legs hanging and knees at 90°), the length is recorded to the nearest millimeter from the right inguinal crease to the distal end of the right femur. A mark is placed at the midpoint to define where thigh circumference will be measured. |
| **Body mass** | Weight (kg) | Weight is measured on a Toledo digital scale with the subject standing still in the center of the scale platform facing the recorder, hands at side, and looking straight ahead. The measurement is recorded in pounds and then automatically converted to kilograms. |
| | Body mass index (kg/$m^2$) | Calculated by dividing the body weight (in kilograms) by the squared body height (in meters). |

| | | |
|---|---|---|
| **Visceral adiposity** | Waist circumference (cm) | With the subject standing, the measuring tape is placed around the trunk right above the right iliac crest, parallel to the floor. The tape should be snug without compressing the skin. The circumference is recorded at the end of expiration to the nearest millimeter. |
| | Wast-to-height ratio | Calculated by dividing the waist circumference by the body height (both in centimeters). |
| **Subcutaneous adiposity** | Triceps skinfold (mm) | With the subject standing (shoulders relaxed, arms hanging freely), enough skin and adipose tissue is grasped to form a distinct fold that separates from the underlying muscle about two centimeters above the midpoint of the posterior surface of the arm (previously marked). The skinfold thickness is recorded to the nearest 0.1 mm with a caliper. |
| | Subscapular skinfold (mm) | With the subject standing (shoulders relaxed, arms hanging freely), a fold of skin and adipose tissue is grasped right above and medial to the inferior angle of the right scapula. The skinfold should form a line of about 45 degrees extending towards the right elbow and its thickness is recorded to the nearest 0.1 mm with a caliper. |
| **Primarily lean mass** | Mid-upper arm circumference (cm) | With the subject standing (shoulders relaxed, right arm hanging loosely without tightening the muscles), the measuring tape is placed around the midpoint of the upper arm (previously marked) perpendicular to its long axis. The two ends of the tape are pulled together without compressing the skin and circumference is measured to the nearest millimeter. |
| | Mid-thigh circumference (cm) | With the subject standing (right leg forward, knee slightly flexed), the measuring tape is placed around the midpoint of the upper leg (previously marked) perpendicular to its long axis. The two ends of the tape are pulled together without compressing the skin and circumference is measured to the nearest millimeter. |

**SUPPLEMENTARY TABLE 2.** Sociodemographic characteristics, medical history, mortality data and anthropometric measurements of participants aged ≥20 years from NHANES-III and NHANES-IV used in our study. Anthropometric variables are stratified by sex (F: Females, M: Males). Continuous variables are presented as median (interquartile range) and categorical variables as absolute frequency (%).

| Characteristics | Overall (n=18,794) | NHANES-III (n=11,774) | NHANES-IV (n=7,020) | P-value |
|---|---|---|---|---|
| **Sociodemographic** | | | | |
| Female (%) | 9505 (50.6) | 6046 (51.4) | 3459 (49.3) | 0.006 |
| Age (years) | 45 (31-64) | 44 (31-64) | 46 (31-63) | **0.408** |
| PhenoAge (years) | 56.8 (42.59-76.2) | 57.95 (43.79-78.14) | 55.05 (40.56-72.61) | <0.001 |
| Non-Hispanic White (%) | 8761 (46.6) | 5124 (43.5) | 3637 (51.8) | |
| Non-Hispanic Black (%) | 4246 (22.6) | 3043 (25.8) | 1203 (17.1) | |
| Mexican-American (%) | 4760 (25.3) | 3172 (26.9) | 1588 (22.6) | <0.001 |
| **Medical history** | | | | |
| ≥1 comorbidity (%) | 9268 (49.3) | 5614 (47.7) | 3654 (52.1) | <0.001 |
| Hypertension (%) | 4996 (26.6) | 3040 (25.8) | 1956 (27.9) | **0.568** |
| Arthritis (%) | 4124 (21.9) | 2473 (21) | 1651 (23.5) | 0.002 |
| Asthma (%) | 1583 (8.4) | 866 (7.4) | 786 (11.2) | <0.001 |
| Diabetes (%) | 1399 (7.4) | 797 (6.8) | 545 (7.8) | <0.001 |
| Bronchitis (%) | 1036 (5.5) | 659 (5.6) | 533 (7.6) | 0.004 |
| Malignancy (%) | 987 (5.3) | 539 (4.6) | 377 (5.4) | **0.111** |
| Heart attack (%) | 825 (4.4) | 442 (3.8) | 286 (4.1) | 0.029 |
| Heart failure (%) | 606 (3.2) | 414 (3.5) | 209 (3) | **0.929** |
| Stroke (%) | 496 (2.6) | 287 (2.4) | 192 (2.7) | **0.531** |
| Emphysema (%) | 373 (2) | 235 (2) | 138 (2) | <0.001 |
| **Mortality data** | | | | |
| Mortality (%) | 5853 (31.1) | 4545 (38.6) | 1308 (18.6) | <0.001 |
| Follow-up (months) | 191 (131-288) | 274 (182-302) | 139 (121-155) | <0.001 |

| Characteristics | | Overall (n=18,794) | NHANES-III (n=11,774) | NHANES-IV (n=7,020) | P-value |
|---|---|---|---|---|---|
| **Anthropometric measurements** | | | | | |
| Height (cm) | F | 160.3 (155.7-165.4) | 160 (155.5-165.1) | 160.9 (156-165.9) | <0.001 |
| | M | 173.7 (168.7-179) | 173.3 (168.5-178.4) | 174.5 (169.2-180) | <0.001 |
| Arm length (cm) | F | 34.8 (33.4-36.3) | 34.5 (33-36) | 35.5 (34-37) | <0.001 |
| | M | 38 (36.5-39.5) | 37.5 (36-39) | 38.7 (37.2-40.3) | <0.001 |
| Leg length (cm) | F | 38.3 (36.1-40.5) | 38.3 (36.1-40.6) | 38.2 (36.1-40.3) | **0.081** |
| | M | 42 (39.8-44.3) | 42 (39.7-44.2) | 42.1 (40-44.4) | 0.013 |
| Weight (kg) | F | 66.35 (57.85-76.55) | 65.8 (57.4-76.69) | 67.3 (58.5-76.35) | 0.008 |
| | M | 78.6 (69.6-88.7) | 77.6 (68.65-87.61) | 80.2 (71.2-90.5) | <0.001 |
| BMI (kg/m2) | F | 25.85 (22.6-29.7) | 25.8 (22.4-29.8) | 25.92 (22.88-29.48) | **0.473** |
| | M | 26.07 (23.47-28.94) | 25.8 (23.3-28.7) | 26.43 (23.85-29.26) | <0.001 |
| Waist circumference (cm) | F | 89.2 (80-99) | 88.8 (79-98.8) | 89.9 (81.25-99.3) | <0.001 |
| | M | 95 (86.7-103.3) | 94.2 (85.9-102.6) | 96.3 (88.3-104.5) | <0.001 |
| Waist-to-height ratio | F | 0.56 (0.5-0.62) | 0.56 (0.49-0.62) | 0.56 (0.5-0.62) | 0.002 |
| | M | 0.55 (0.5-0.6) | 0.54 (0.49-0.59) | 0.55 (0.5-0.6) | <0.001 |
| Subscapular skinfold (cm) | F | 21.2 (14.7-28.4) | 21.9 (14.83-29.6) | 20.2 (14.5-26.2) | <0.001 |
| | M | 18.5 (13.3-24) | 18.5 (13.2-24) | 18.4 (13.6-23.6) | **0.969** |
| Triceps skinfold (mm) | F | 22.9 (17.2-29.1) | 23 (16.9-29.7) | 22.8 (17.4-28.4) | 0.031 |
| | M | 11.7 (8.7-15.4) | 11.25 (8.3-14.9) | 12.4 (9.3-16.4) | <0.001 |
| Arm circumference (cm) | F | 30 (27.4-33.2) | 30.1 (27.4-33.5) | 29.9 (27.4-32.7) | <0.001 |
| | M | 32.4 (30.1-34.9) | 32.3 (30-34.7) | 32.6 (30.3-35.1) | <0.001 |
| Thigh circumference (cm) | F | 50 (46.2-54.6) | 49.9 (46.02-54.5) | 50.1 (46.6-54.7) | 0.048 |
| | M | 50.9 (47.5-54.3) | 50.4 (47.1-53.8) | 51.6 (48.2-55.2) | <0.001 |

**SUPPLEMENTARY TABLE 3.** Gompertz proportional hazard regression models for AnthropoAge. All models were fitted to predict 10-year all-cause mortality in NHANES-III using anthropometric measurements stratified by sex, with ethnicity in the shape parameter. Transformed variables and orthogonal polynomials (OP) were used as described in methods. Model selection was conducted by BIC minimization. No variable presented multicollinearity assessed with the variance inflation factor.

| Model | Parameter | β-coefficient | Lower 95%CI | Upper 95%CI |
|---|---|---|---|---|
| **AnthropoAge Males BIC 31734.5** | Shape | 0.0062 | 0.0055 | 0.0068 |
| | Rate | -7.2887 | -8.6857 | -5.8917 |
| | Chronological Age | 0.0705 | 0.0670 | 0.0739 |
| | WHtR (OP,1) | 14.8761 | 9.5059 | 20.2464 |
| | WHtR (OP,2) | 5.4062 | 2.4693 | 8.3431 |
| | Arm circumference | -0.6453 | -0.8839 | -0.4068 |
| | Thigh circumference (OP,1) | -6.7501 | -12.8232 | -0.6770 |
| | Thigh circumference (OP,2) | 4.2013 | 1.7448 | 6.6578 |
| | Shape (Non-Hispanic Black) | 0.0010 | 0.0004 | 0.0017 |
| | Shape (Mexican-American) | -0.0004 | -0.0011 | 0.0002 |
| | Shape (Other race/ethnicities) | -0.0042 | -0.0049 | -0.0036 |
| **AnthropoAge Females BIC 27743.2** | Shape | 0.0079 | 0.0072 | 0.0072 |
| | Rate | -20.0987 | -21.9140 | -21.9140 |
| | Chronological Age | 0.0770 | 0.0734 | 0.0734 |
| | Weight | 1.1911 | 0.7006 | 0.7006 |
| | WHtR | 6.4819 | 4.6758 | 4.6758 |
| | Subscapular skinfold | -0.3473 | -0.5415 | -0.5415 |
| | Triceps skinfold | -0.4526 | -0.6814 | -0.6814 |
| | Thigh circumference (OP,1) | -16.3648 | -23.0983 | -23.0983 |
| | Thigh circumference (OP,2) | 7.5900 | 4.8101 | 4.8101 |
| | Shape (Non-Hispanic Black) | 0.0003 | -0.0004 | -0.0004 |
| | Shape (Mexican-American) | -0.0011 | -0.0018 | -0.0018 |
| | Shape (Other race/ethnicities) | -0.0024 | -0.0031 | -0.0031 |

**SUPPLEMENTARY TABLE 4.** Gompertz proportional hazard regression models for S-AnthropoAge. All models were fitted to predict 10-year all-cause mortality in NHANES-III using transformed body-mass index (BMI) and waist to height ratio (WHtR) stratified by sex, with ethnicity in the shape parameter. Orthogonal polynomials (OP) were used as described in methods. Model selection was conducted by BIC minimization. No variable presented multicollinearity assessed with the variance inflation factor.

| Model | Parameter | β - coefficient | Lower 95%CI | Upper 95%CI |
|---|---|---|---|---|
| **S-AnthropoAge Males BIC 31720** | Shape | 0.0060 | 0.0053 | 0.0068 |
| | Rate | -19.0818 | -21.3185 | -16.8451 |
| | Chronological Age | 0.0733 | 0.0702 | 0.0765 |
| | BMI (OP,1) | -26.6759 | -33.6266 | -19.7252 |
| | BMI (OP,2) | 12.3235 | 9.5685 | 15.0785 |
| | WHtR | 9.7851 | 6.9656 | 12.6046 |
| | Shape (Non-Hispanic Black) | 0.0010 | 0.0002 | 0.0018 |
| | Shape (Mexican American) | -0.0001 | -0.0009 | 0.0007 |
| | Shape (Other race/ethnicities) | -0.0044 | -0.0052 | -0.0036 |
| **S-AnthropoAge Females BIC 27808.2** | Shape | 0.0077 | 0.0069 | 0.0069 |
| | Rate | -19.2580 | -21.1427 | -21.1427 |
| | Chronological Age | 0.0818 | 0.0784 | 0.0784 |
| | BMI (OP,1) | -20.8035 | -28.0811 | -28.0811 |
| | BMI (OP,2) | 9.2458 | 6.3241 | 6.3241 |
| | WHtR | 8.5259 | 6.1679 | 6.1679 |
| | Shape (Non-Hispanic Black) | 0.0004 | -0.0004 | -0.0004 |
| | Shape (Mexican American) | -0.0008 | -0.0016 | -0.0016 |
| | Shape (Other race/ethnicities) | -0.0025 | -0.0033 | -0.0033 |

**SUPPLEMENTARY TABLE 5.** Area under the receiving operating characteristic curve (AUROC) with 95% confidence intervals for AnthropoAge, S-AnthropoAge and PhenoAge stratified by sex and number of comorbidities. There were no significant differences between the three metrics in any category when performing ROC tests with the *pROC* R package. Comorbidities assessed included: diabetes, hypertension, asthma, arthritis, heart failure, history of heart attack or stroke, emphysema, bronchitis, and malignant neoplasms.

| Strata | | AnthropoAge | S-AnthropoAge | PhenoAge |
|---|---|---|---|---|
| **Sex** | Men | 0.876 (0.861, 0.892) | 0.876 (0.861, 0.892) | 0.874 (0.859, 0.890) |
| | Women | 0.879 (0.862, 0.896) | 0.874 (0.857, 0.892) | 0.881 (0.864, 0.898) |
| **Comorbidities** | 0 | 0.810 (0.773, 0.847) | 0.809 (0.772, 0.846) | 0.803 (0.766, 0.840) |
| | 1 | 0.848 (0.825, 0.871) | 0.846 (0.822, 0.869) | 0.849 (0.826, 0.872) |
| | ≥2 | 0.816 (0.795, 0.836) | 0.814 (0.793, 0.835) | 0.830 (0.810, 0.850) |

**SUPPLEMENTARY TABLE 6.** Hazard ratios (HR) with 95% confidence intervals and standard errors (SE) for cause-specific and all-cause mortality using AnthropoAge, S-AnthropoAge and PhenoAge. The highest HR for each mortality cause is highlighted in bold. All models are adjusted for sex, number of comorbidities and race/ethnicity.

| Mortality | AnthropoAge | S-AnthropoAge | PhenoAge |
|---|---|---|---|
| **Cardiovascular** | HR=1.061 (1.058-1.065), SE=0.0018 | **HR=1.062 (1.058-1.066), SE=0.0018** | HR=1.054 (1.051-1.057), SE=0.0014 |
| **Diabetes Mellitus** | HR=1.042 (1.034-1.051), SE=0.0042 | HR=1.043 (1.034-1.051), SE=0.0041 | **HR=1.055 (1.047-1.063), SE=0.004** |
| **Stroke** | HR=1.069 (1.062-1.076), SE=0.0034 | **HR=1.069 (1.062-1.077), SE=0.0035** | HR=1.055 (1.05-1.061), SE=0.0026 |
| **Cancer** | **HR=1.043 (1.04-1.046), SE=0.0015** | HR=1.043 (1.04-1.046), SE=0.0015 | HR=1.039 (1.036-1.041), SE=0.0013 |
| **Influenza/ Pneumonia** | **HR=1.076 (1.064-1.089), SE=0.0058** | HR=1.076 (1.063-1.088), SE=0.0058 | HR=1.064 (1.055-1.074), SE=0.0045 |
| **Nephritis/ Nephrosis** | HR=1.061 (1.045-1.076), SE=0.0074 | HR=1.061 (1.046-1.077), SE=0.0075 | **HR=1.064 (1.052-1.076), SE=0.0058** |
| **Alzheimer** | HR=1.081 (1.072-1.09), SE=0.0042 | **HR=1.083 (1.074-1.092), SE=0.0041** | HR=1.061 (1.055-1.067), SE=0.0029 |
| **Chronic lower respiratory** | HR=1.053 (1.045-1.060), SE=0.0038 | **HR=1.054 (1.046-1.062), SE=0.0039** | HR=1.041 (1.035-1.047), SE=0.0029 |
| **All-cause mortality** | **HR=1.081 (1.078-1.083), SE=0.001** | HR=1.080 (1.078-1.083), SE=0.001 | HR=1.069 (1.067-1.070), SE=0.0008 |

### SUPPLEMENTARY FIGURES

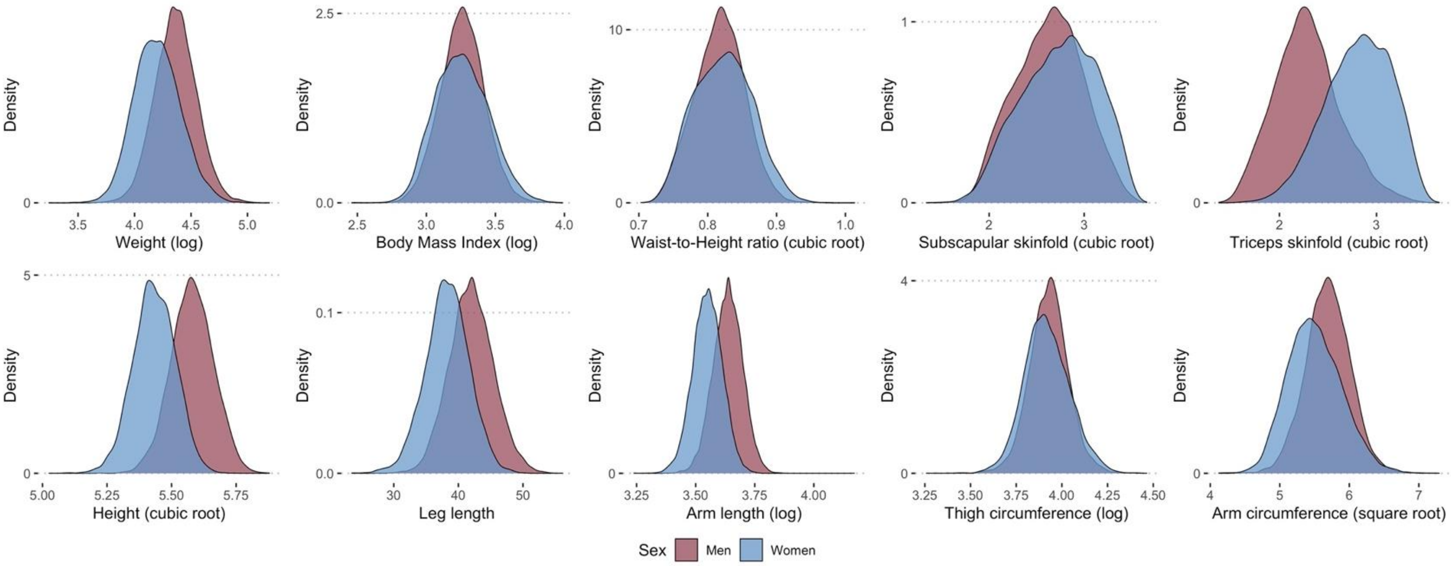


| Variable | A-statistic (prior) | p-value (prior) | Transformation | A-statistic (transformed) | p-value (transformed) |
|---|---|---|---|---|---|
| Weight (kg) | 68.117 | 3.7e-24 | Logarithmic | 1.791 | 1.403e-4 |
| Body Mass Index (kg/m2) | 88.739 | 3.7e-24 | Logarithmic | 3.864 | 1.264e-9 |
| Waist-to-Height ratio | 26.634 | 3.7e-24 | Cubic root | 7.171 | 1.573e-17 |
| Subscapular skinfold (cm) | 112.876 | 3.7e-24 | Cubic root | 32.330 | 3.7e-24 |
| Triceps skinfold (cm) | 238.069 | 3.7e-24 | Cubic root | 52.236 | 3.7e-24 |
| Height (cm) | 14.563 | 3.7e-24 | Cubic root | 11.474 | 3.7e-24 |
| Leg length (cm) | 2.052 | 3.225e-5 | None | 2.052 | 3.225e-5 |
| Arm length (cm) | 13.289 | 3.7e-24 | Logarithmic | 6.696 | 2.098e-16 |
| Thigh circumference (cm) | 52.069 | 3.7e-24 | Logarithmic | 8.719 | 3.618e-21 |
| Arm circumference (cm) | 8.106 | 9.87e-20 | Square root | 1.416 | 1.167e-3 |

**Supplementary Figure 1.** Distribution of anthropometric variables after transformation, stratified by sex. We also provide a table specifying the transformation that was performed for each variable, as well as the A-statistic and p-value from the Anderson-Darling test prior and after transformation. These transformations were the ones we used for the development of AnthropoAge and S-AnthropoAge.

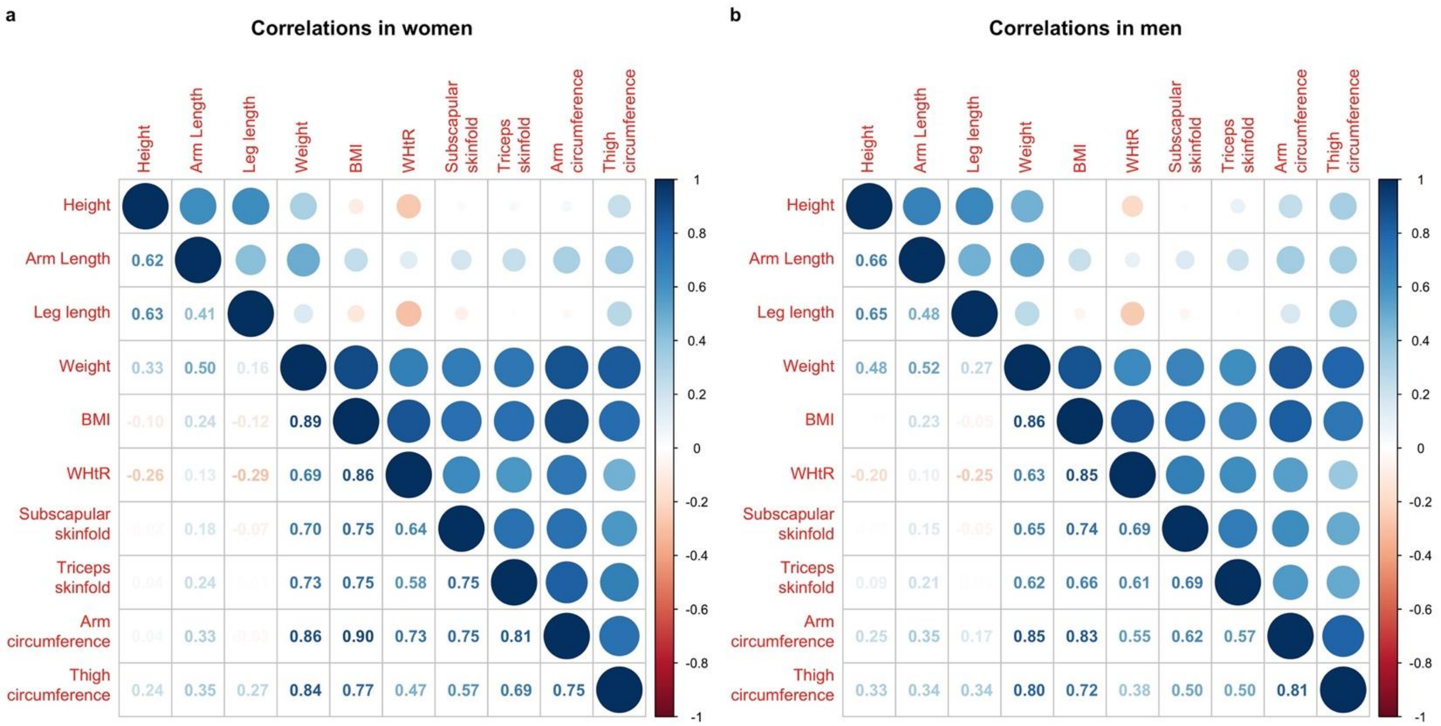


**Supplementary Figure 2.** Linear correlations between included anthropometric measurements calculated separately for women (a) and men (b).

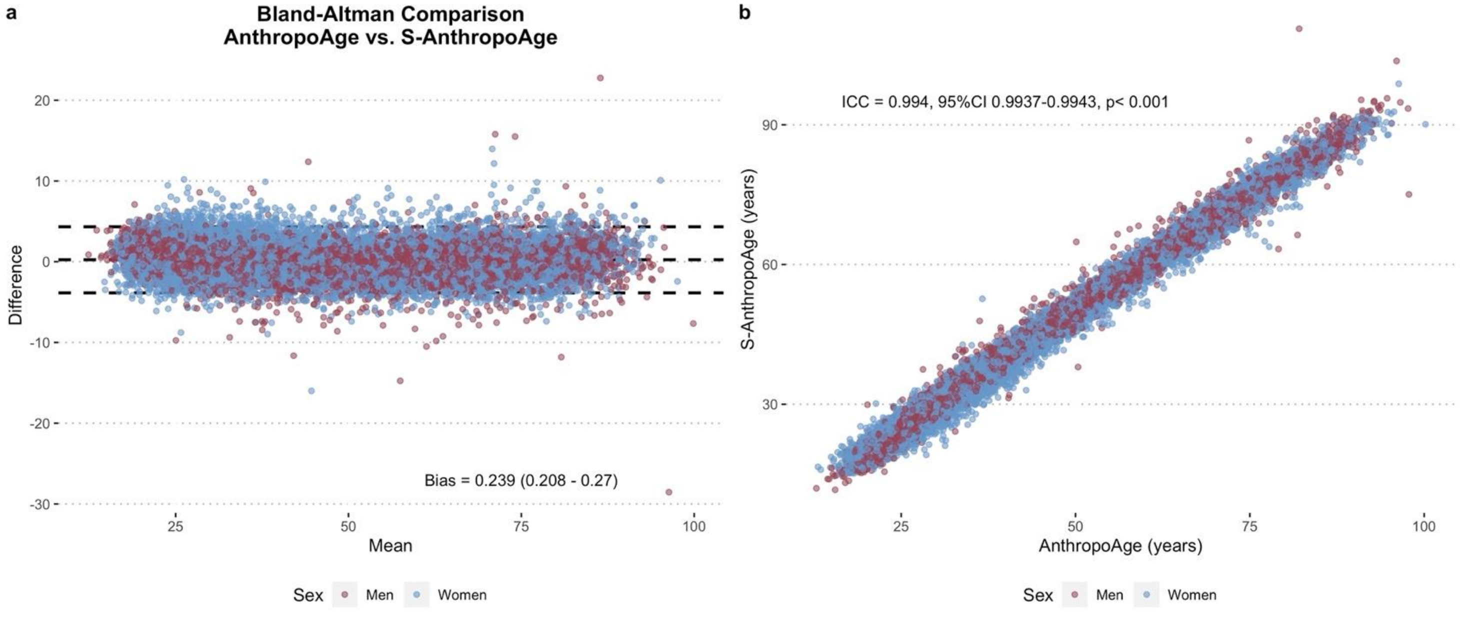


**Supplementary Figure 3.** Bland-Altman plot comparing AnthropoAge and S-AnthropoAge estimations (**A**) and scatterplot of AnthropoAge vs. S-AnthropoAge (**B**), stratified by sex. The *blandr* R package was used for this analysis (https://doi.org/10.5281/zenodo.824514).

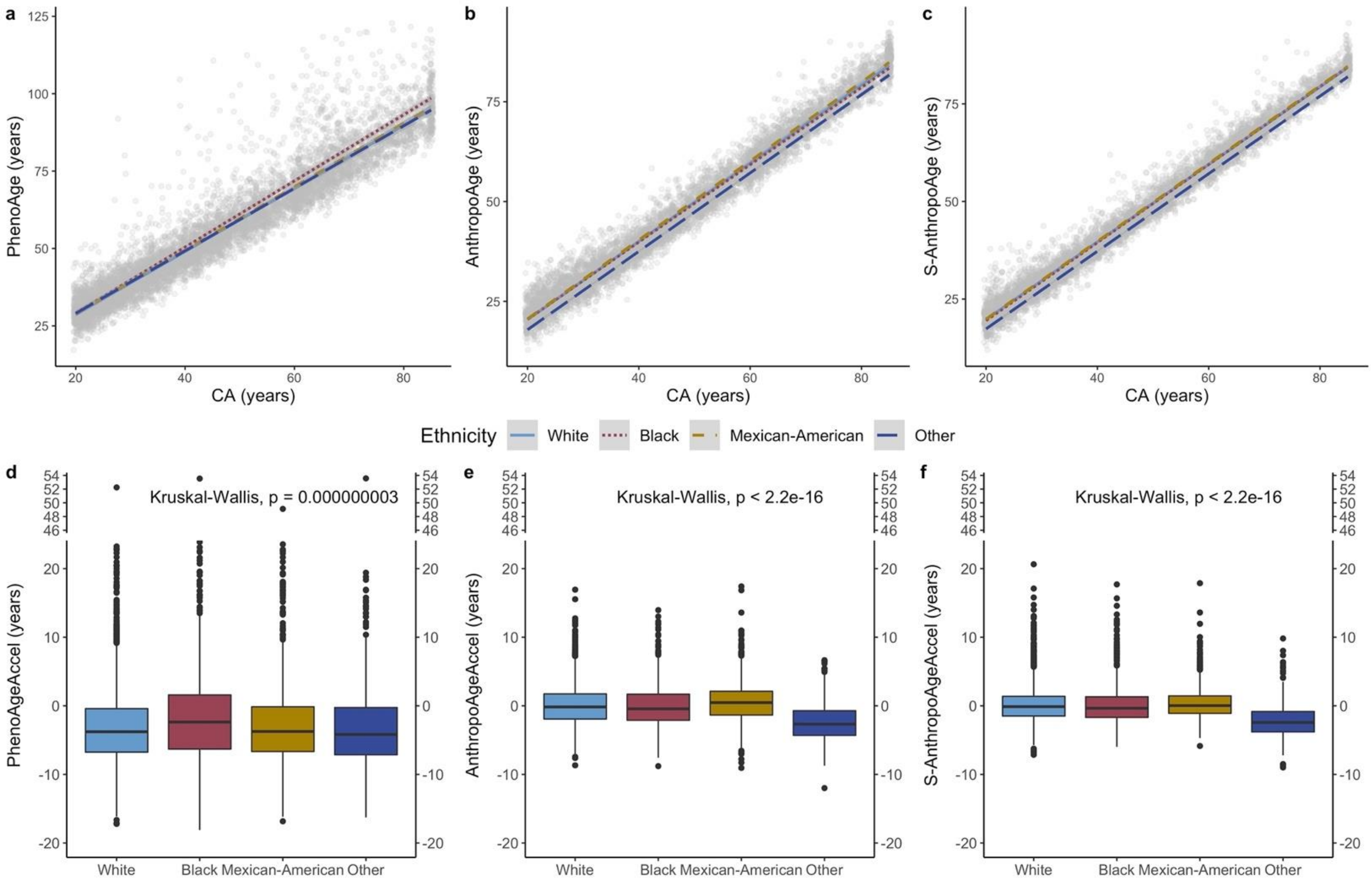


**Supplementary Figure 4.** Scatter plots of CA vs PhenoAge, AnthropoAge and S-AnthropoAge (a-c) and boxplots of accelerated metrics (d-f) compared across different races/ethnicities in the validation cohort. Median PhenoAgeAccel: -3.78 years (White), **-2.36 (Black),** -3.74 (Mexican-American), -4.18 (Other). Median AnthropoAgeAccel: -0.16 (White), -0.44 (Black), **0.47 (Mexican-American)**, -2.67 (Other). Median S-AnthropoAgeAccel: -0.12 (White), -0.35 (Black), **0.04 (Mexican-American),** -2.42 (Other). *ggbreak* R package was used to produce axis breaks (https://doi.org/10.3389/fgene.2021.774846).

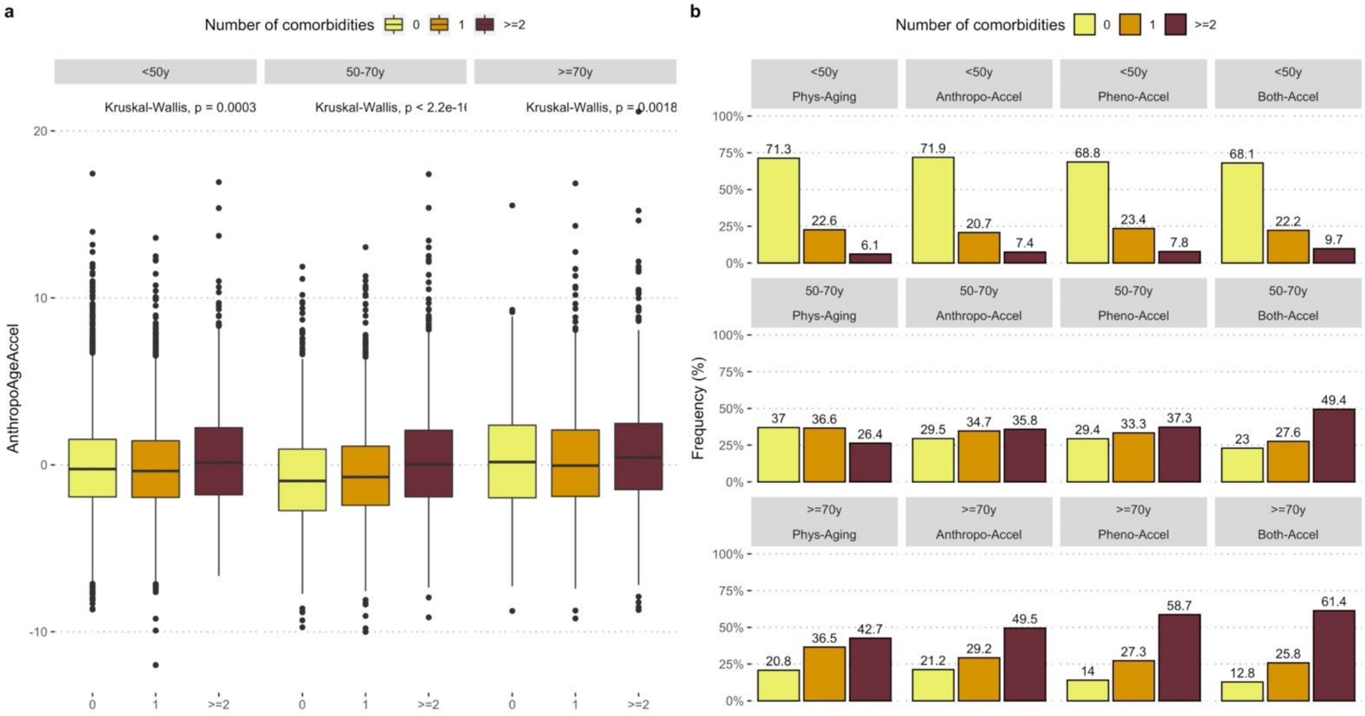


**Supplementary Figure 5.** Comparison of AnthropoAgeAccel across number of comorbidities stratified by age (a). We also show a comparison of multimorbidity rates between physiological aging, AnthropoAge or PhenoAge acceleration and multidomain acceleration (b).

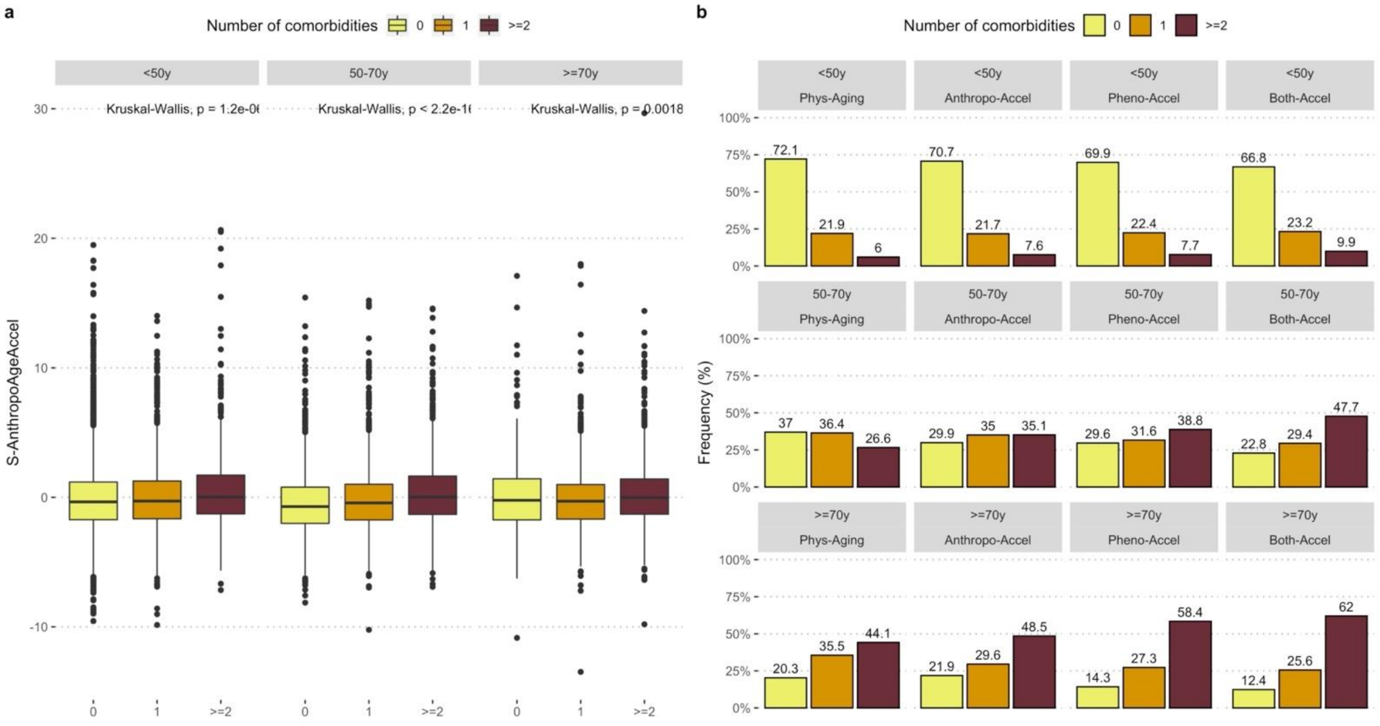


**Supplementary Figure 6.** Same as above but assessing S-AnthropoAgeAccel instead.

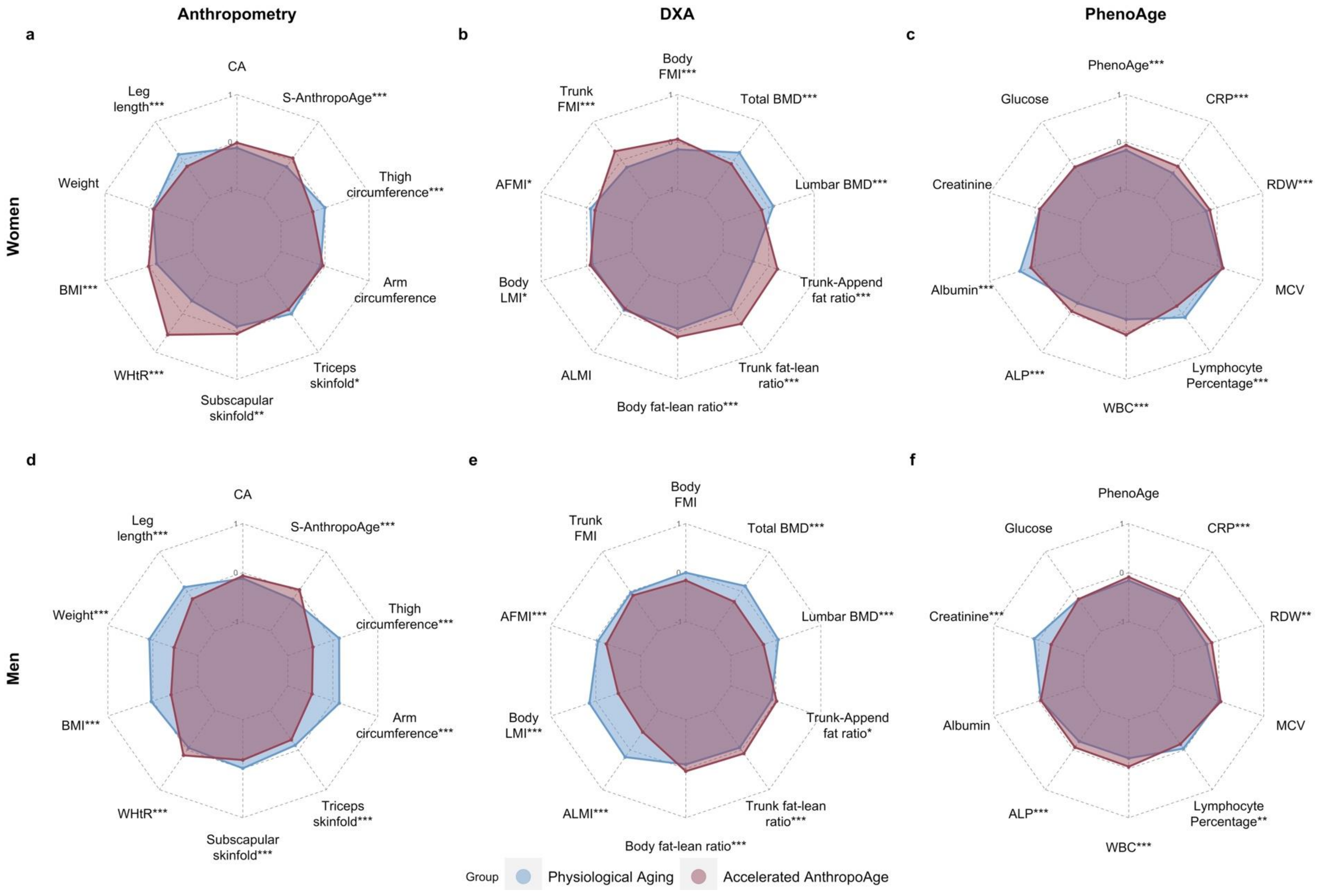


**Supplementary Figure 7.** Spider plots stratified by sex comparing patterns of body composition and PhenoAge components across cases with physiological (S-AnthropoAgeAccel ≤0) and accelerated aging (S-AnthropoAgeAccel >0). Variables were scaled and compared using Mann-Whitney U test (p-value <0.05: *, <0.01: **, <0.001: ***). Spider plots were generated using the *fmsb* R package (https://CRAN.R-project.org/package=fmsb).

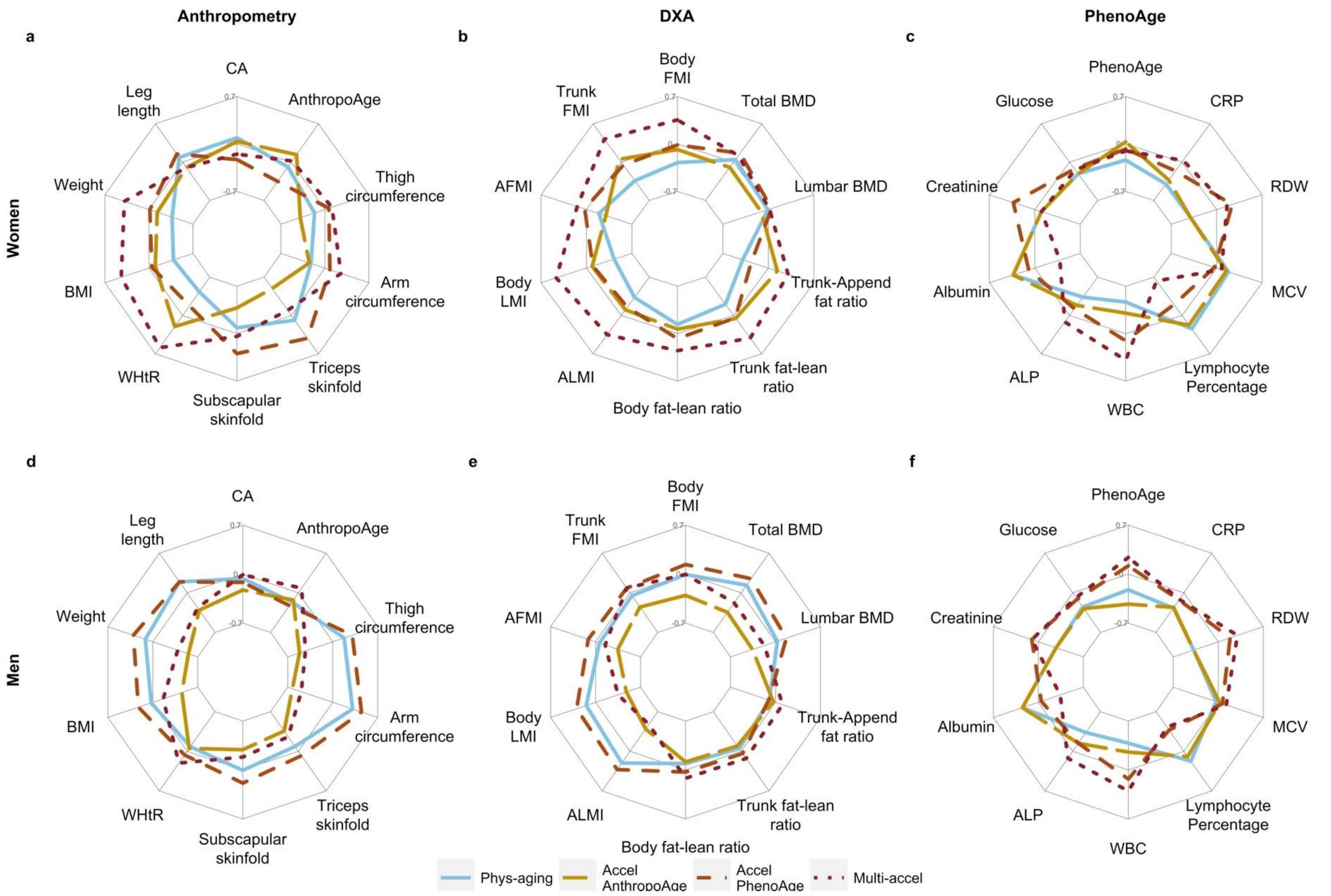


**Supplementary Figure 8.** Spider plots stratified by sex comparing patterns of body composition and PhenoAge components across subjects using the multidomain aging indicator in the following categories: physiological aging, accelerated AnthropoAge, accelerated PhenoAge and multidomain acceleration.

<u>Abbreviations</u>: AFMI: Appendicular Fat Mass Index. ALMI: Appendicular Lean Mass Index. ALP: Alkaline Phosphatase. BMD: Bone Mineral Density. BMI: Body Mass Index. CA: Chronological Age. CRP: C-Reactive Protein. MCV: Mean Corpuscular Volume. RDW: Red blood cell Distribution Width. WBC: White Blood Cell count. WHtR: Waist-to-Height-ratio
